# The long-term risk of tuberculosis among individuals with Xpert Ultra “trace” screening results: a longitudinal follow-up study

**DOI:** 10.1101/2025.03.20.25324205

**Authors:** Joowhan Sung, Mariam Nantale, Annet Nalutaaya, Patrick Biché, James Mukiibi, Joab Akampurira, Rogers Kiyonga, Francis Kayondo, Michael Mukiibi, Caitlin Visek, Caleb E Kamoga, David W Dowdy, Achilles Katamba, Emily A Kendall

## Abstract

**Background:** Systematic screening for tuberculosis using Xpert Ultra generates “trace” results of uncertain significance. Additional microbiological testing in this context is often negative, but untreated individuals might still progress to culture-positive disease. We aimed to estimate the two-year risk of tuberculosis among screening participants with trace-positive sputum.

**Methods:** We screened 31,505 people for tuberculosis in Uganda using sputum Xpert Ultra as an initial test, through event-based and door-to-door screening. We enrolled 128 participants with trace-positive sputum (PWTS), 139 Ultra-negative controls into a prospective cohort, and 110 Ultra-positive (>trace) controls for cross-sectional comparison. All participants underwent extensive initial evaluation, and untreated PWTS and negative controls were followed with re-testing for up to 24 months. We estimated cumulative hazards of tuberculosis among PWTS versus negative controls, using two definitions of tuberculosis: one incorporating clinician judgment (primary) and one based strictly on microbiological results (secondary). We then compared hazards between subgroups of PWTS.

**Findings:** Of 128 PWTS, 79 (62%) were male and 19 (15%) HIV-positive. Forty-five (35%) PWTS were recommended for treatment upon enrollment, eight lost to follow-up within three months, and 75 followed for median 706 (interquartile range 344-714) days, of whom 19 were recommended for treatment during follow-up. The cumulative hazard of tuberculosis among PWTS not treated at baseline was 0.24 (95% confidence interval: 0.15-0.40) at one year and 0.33 (0.21-0.54) at two years, versus 0.03 (0.01-0.10) at two years for negative controls. Hazards were similar for microbiologically defined tuberculosis (0.36 [0.22-0.58] at 2 years). Tuberculosis diagnosis during follow-up was strongly associated with abnormal baseline chest X-ray (hazard ratio 14.6 [3.3-63.8]) but not with baseline symptoms.

**Interpretation:** Individuals with trace-positive sputum during screening have a substantial two-year risk of tuberculosis, even when extensive initial evaluations do not confirm disease. Treatment should be considered for most screening participants with trace-positive sputum and abnormal chest imaging.

**Funding:** National Institutes of Health and Gates Foundation

**Research in context:** *Evidence before this study:* Recent advances in tuberculosis research have shifted the disease framework from a binary classification of latent versus active tuberculosis to a continuum of disease states. They have also led to a better understanding of the dynamic disease course of early tuberculosis, which can either progress to culture-positive disease or regress spontaneously over time. “Trace” results from Xpert MTB/RIF Ultra (“Ultra”) are sometimes perceived as false positives in individuals who subsequently test negative on additional diagnostic assays. However, some of these individuals may have early tuberculosis that falls below the detection threshold of existing diagnostic tests and could progress to microbiologically detectable disease over time. In screening contexts, trace-positive Ultra results constitute up to half of all positive results, and it is critical to understand how to interpret this result from screening participants, particularly among individuals without further evidence of tuberculosis on additional testing. To investigate this, we searched PubMed for studies published in English from inception to up to February 7, 2025, using the terms “tuberculosis” AND (“Xpert” OR “Xpert Ultra” OR “Ultra”) AND “Trace” and also reviewed the reference lists of relevant search results. In two prevalence surveys that each used a combination of symptom and chest X-ray as the screening test with Ultra as a confirmatory test, the proportion with positive cultures among those with trace-positive sputum ranged from 20% to 38%. In a study conducted in Uganda where Ultra was used as an initial screening test, only 14% of individuals with a trace-positive result had positive sputum cultures. However, prior studies were limited to a one-time evaluation of individuals following a trace result. None longitudinally followed these individuals after a negative initial work-up to assess their long-term risk of tuberculosis and its predictors, and appropriate management strategy for such individuals remains unknown.

*Added value of this study:* In this study, individuals with Ultra trace-positive screening results who were not started on treatment after extensive diagnostic testing were followed for up to two years with repeated testing. The 2-year cumulative hazard of tuberculosis disease was substantial at 0.33 (95% confidence interval 0.21-0.54), compared to 0.03 (0.01-0.10) among individuals with Ultra-negative screening results. Those who had a normal chest X-ray at enrollment were at significantly lower risk of developing tuberculosis. Two-year tuberculosis risk was similar between those who reported symptoms at the time of enrollment and those who did not.

*Implications of all the available evidence:* The high two-year risk of tuberculosis observed among people with trace results in this study, even when Ultra was used without any prior screening step, support provision of treatment for tuberculosis disease to most individuals who receive trace results during tuberculosis screening interventions. These results also demonstrate that X-ray could be a useful tool to guide treatment decision-making for individuals with trace-positive sputum.

## Introduction

Despite being a curable disease, tuberculosis continues to be the leading single-agent infectious cause of death worldwide, accounting for 1.25 million deaths in 2023.^1^ About 25% of individuals who develop tuberculosis are never diagnosed and started on treatment ^1^. Detecting tuberculosis at early stages through effective diagnostic strategies could reduce transmission and prevent severe outcomes.

Tuberculosis programs are increasingly conducting community-based tuberculosis screening,^2^ and during such screening efforts, Xpert MTB/RIF Ultra (“Ultra”; Cepheid), a molecular diagnostic tool for tuberculosis, is widely used as a confirmatory test for individuals with tuberculosis symptoms and/or abnormal chest X-rays. In screening contexts, a large proportion of positive Ultra results are in the “trace” category.^3–5^ Trace results, which represent the lowest level of *M. tuberculosis (Mtb)* DNA detection, are reported when the Ultra assay detects one of its multicopy amplification targets (*IS1660* or *IS1810*) but not its single-copy *rpoB* gene target.^6^ Studies that have used Ultra as an initial or confirmatory test during general-population screening in high-burden settings have reported that 24% to 46% of positive results are trace-positive.^3–5, 7^

Due to lower tuberculosis prevalence among general populations than compared to symptomatic patients tested at health facilities, the positive predictive value of a trace result could be lower in screening contexts, but it is uncertain what proportion of trace results reflect underlying tuberculosis disease. We previously reported on systematic screening for tuberculosis that we conducted in Uganda, using Ultra as an initial test regardless of symptoms, with detailed baseline evaluation of participants who received trace results.^5^ Among the first 92 such participants, only 22 (24%) had microbiologically confirmed tuberculosis, with just 13 (14%) having a positive culture. However, tuberculosis has a dynamic and varied disease course, with progression and regression of bacterial burden occurring across different states of infection. ^8–10^ We hypothesized that some individuals with trace-positive sputum who are otherwise microbiologically negative may have early (or otherwise minimal-burden) disease that could progress over time, such that they could potentially benefit from preventive or empirical treatment. No existing studies have evaluated the long-term risk of tuberculosis among individuals with trace-positive sputum who otherwise do not have current evidence for tuberculosis. Therefore, within the same cohort after additional enrollment, we aimed to estimate the two-year risk of tuberculosis disease among participants with trace-positive screening results who had completed detailed evaluations for tuberculosis without definite evidence of tuberculosis.

## Methods

### Symptom-Neutral Tuberculosis Screening and Participant Recruitment

We conducted Ultra-based systematic screening for tuberculosis in Kampala, Uganda, from February 2021 to April 2024, enrolling participants with trace-positive sputum (PWTS) as well as Ultra-positive and Ultra-negative controls. Procedures for recruitment, described previously,^5^ were continued through April 2024 and are detailed in appendix (**p 3**). Briefly, participants for tuberculosis screening were recruited primarily through community-based screening events and door-to-door screening and to a lesser extent through contact investigations. Ultra sputum testing was offered to any individuals aged ≥15 years who were not on active tuberculosis treatment, regardless of their symptoms. Upon receiving the Ultra results from the study’s screening activities, we recruited all PWTS, as well as age- and sex-matched participants with negative screening results (“negative controls”) and consecutive participants with positive (>trace) screening results (“positive controls”), for further diagnostic evaluation (**appendix p 4)**. To enhance our sample size, we also enrolled people who were found to have trace-positive sputum during community-based, symptom-agnostic Ultra testing that was offered in high-risk areas of Kampala during a national screening campaign.^11^ Tuberculosis screening continued until 128 PWTS were enrolled (sample size explained in **appendix pp 5-6**).

### Evaluation and Follow-Up of Study Participants

Upon enrollment, PWTS received extensive diagnostic testing, including symptom survey (**appendix pp 7-8**), repeat sputum Ultra, two sets of sputum liquid and solid mycobacterial cultures, HIV testing (using a serial rapid testing algorithm,^12^ if not known positive), chest X-ray (CXR), and chest computed tomography (CT). Participants with HIV also received urine lipoarabinomannan (LAM) testing (Determine TB LAM; Abbott). Participants screened after June 2021 were additionally surveyed about tuberculosis symptoms at the time of screening (**appendix p 9**). A panel of experienced Ugandan pulmonologists and radiologists (“consultants”) reviewed all available new results — including clinical risk factors, symptoms, laboratory results, and chest imaging — for PWTS and for negative controls with abnormal findings, and made recommendations for additional diagnostic testing and/or initiation of treatment. PWTS who were not recommended for tuberculosis treatment or declined treatment were followed with scheduled re-evaluations at 1, 3, 6, 12, and 24 months (**Figure 1**), in addition to unscheduled re-evaluations as needed. At each scheduled follow-up visit, participants were assessed for symptoms and asked to provide expectorated sputum for repeat testing by Ultra. CXRs were repeated at 3 and 12 months, while sputum cultures were repeated for all PWTS at 3 and 12 months, and additionally at 6 and 24 months for PWTS who completed visits after a November 2023 protocol revision. Both negative and positive control-participants underwent the same diagnostic evaluations at baseline. Negative control-participants not recommended for treatment were followed for up to two years, with symptom assessment at 6 months and repeat sputum testing at 12 and 24 months, except for one negative control-participant with a trace result on repeat Ultra testing who received the same frequent follow-up as PWTS. Positive control-participants were referred for treatment initiation.

**Figure 1.**
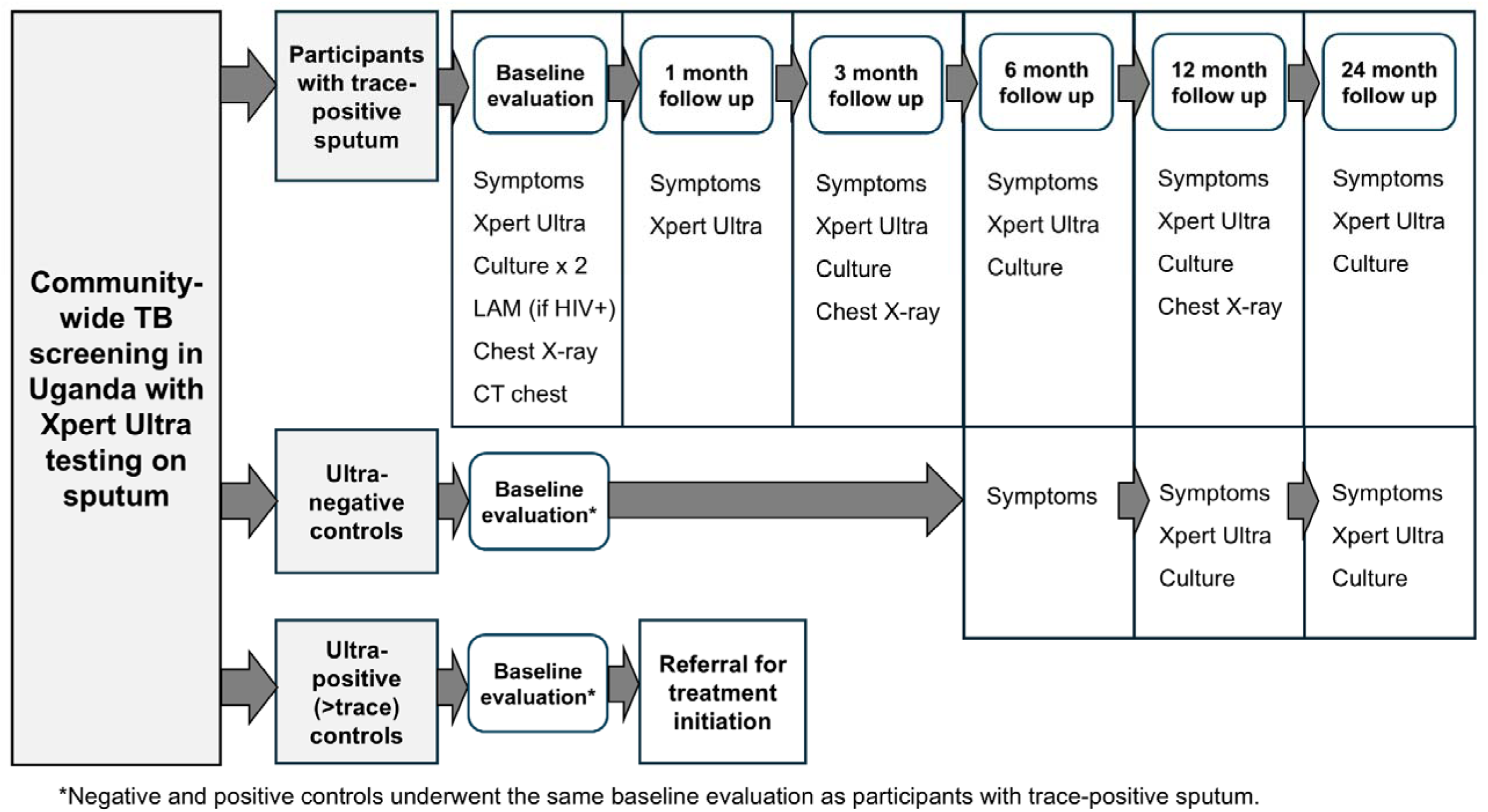
Study procedures for evaluation and follow-up after a trace Ultra result during community-wide screening. Community-based tuberculosis screening was conducted in Kampala, Uganda, from February 2021 to April 2024, using Ultra as the initial test. Participants with a trace-positive result were enrolled and underwent a detailed assessment, including symptom evaluation, additional microbiological testing (repeat sputum Ultra, two sets of sputum liquid and solid mycobacterial cultures, and urine LAM for people with HIV), chest X-ray, and chest CT. Those not initiated on treatment were followed at 1, 3, 6, 12, and 24 months, with repeat testing as outlined below. Sputum cultures at 6 and 24 months were offered to all participants who completed those visits after a November 2023 protocol revision. Both negative and positive controls underwent the same baseline evaluation. Negative controls were followed up to 24 months. One negative control-participant with a trace result on repeat Xpert testing received similarly frequent follow-up as those with an initial trace result. All study follow-up ended in April 2025 (**appendix p 13**).

### Interpretation of Chest Imaging by Radiologists and CAD Software

CXR and CT images of study participants were reviewed by two independent radiologists, with a third radiologist reviewing images if initial readings were discrepant. The radiologists, blinded to clinical information, rated the consistency with current tuberculosis and (separately) with prior tuberculosis on four-point scales. These scores were averaged across all radiologists for reporting (**Appendix p 10**). In addition, all baseline chest X-rays were retrospectively analyzed using CAD software (qXR v4, Qure.ai, India); CAD results were unavailable during treatment decision-making.

### Statistical Analysis

Our primary analysis defined tuberculosis based on treatment recommendation. Under this definition (chosen as primary for the breadth of evidence it considered; see **Appendix p11**), all participants for whom consultants recommended treatment or an external clinician prescribed treatment were counted as having tuberculosis, including those diagnosed clinically. Participants with positive microbiological results that clinicians or consultants interpreted as false-positive were considered not to have tuberculosis. In secondary analyses, we defined tuberculosis based on microbiological positivity, specifically as having a positive sputum Ultra (required to be higher than trace to diagnose based on Ultra alone), a positive sputum culture for *M. tuberculosis* complex, or (among people with HIV) a positive urine LAM. For survival analyses using microbiological positivity to define tuberculosis outcomes, we censored participants who initiated treatment without microbiological confirmation.

We first estimated the cumulative proportion of all PWTS who were recommended for treatment over the course of the study, with a corresponding 95% Wilson score confidence interval (CI). Participants lost to follow-up before three months were excluded from this analysis.

Then, we used survival analysis to evaluate the risk of tuberculosis diagnosis during follow-up among those not diagnosed at baseline, comparing PWTS to negative controls. Participants were classified as having tuberculosis at baseline if their diagnosis was based on the enrollment evaluation, including positive baseline cultures that resulted weeks later. We estimated the cumulative cause-specific hazards of developing tuberculosis among PWTS versus negative controls using the Nelson-Aalen estimator^13^ (**appendix p 12**). We then estimated hazards ratios and corresponding 95% confidence intervals, comparing the two groups, using a Cox proportional hazards model. We performed this analysis for both definitions of tuberculosis, using the corresponding definition of tuberculosis to also define the cohort without tuberculosis at baseline. A sensitivity analysis repeated the same analyses using only matched pairs (**appendix p 4**).

Using similar methods, we estimated the cumulative hazards of tuberculosis diagnosis during follow-up among subgroups of PWTS with different baseline characteristics, and calculated hazard ratios for baseline risk factors, including male sex, HIV infection, positive symptom status (based on a four-symptom assessment), and history of prior tuberculosis treatment. We additionally estimated hazard ratios comparing those with abnormal versus normal baseline chest imaging by each of several definitions, namely (a) CXR read by radiologists as having any abnormalities, (b) CXR read as suggestive of tuberculosis, (c) CXR with qXR tuberculosis score [“CAD score”] ≥0.2, (d) CXR with CAD score ≥0.5, and (e) CT read as suggestive of tuberculosis. All comparisons between subsets of PWTS were performed for each of two definitions of tuberculosis in parallel, and those with missing data for a given risk factor were excluded.

Finally, we compared baseline demographic, clinical, and radiographic characteristics among the following five groups of participants: 1) positive controls, 2) PWTS recommended for treatment at baseline (“Trace, TB at baseline”), 3) PWTS recommended for treatment during follow-up (“Trace, TB during follow-up”), 4) PWTS not recommended for treatment and followed for at least three months (“Trace, no TB”), and 5) negative controls. The CAD scores of baseline CXRs were compared between five groups using Kruskall-Wallis tests followed by Dunn tests with Bonferroni correction.^14^ We also estimated the area under the curve (AUC) for the CAD score at baseline in diagnosing tuberculosis (at baseline or during follow-up, using two definitions) among PWTS who were either diagnosed or followed for ≥three months. In the AUC analysis using the microbiological outcome, participants with microbiologically unconfirmed tuberculosis were considered tuberculosis-negative. This analysis was conducted after all participants had sufficient time to complete twelve months of follow-up (**appendix p 13**). Statistical significance was determined as a two-sided alpha <0.05. Analyses were performed using Stata version 16.1 and R version 4.3.2.

### Ethics Considerations

Informed consent (or adolescent assent with parental consent) was obtained from all study participants. The Institutional Review Boards of the Johns Hopkins University School of Medicine (IRB00269370) and the Makerere University School of Public Health (Protocol 901) approved the study.

## Results

A total of 31,505 individuals aged ≥15 years consented to tuberculosis screening, including 16,568 recruited through event-based screening, 14,782 through door-to-door screening, and 155 through contact investigation. Of these, 31,321 (>99%) successfully provided sputum samples, and 31,150 had valid Ultra results. Among valid results, 297 (1.0% of 31,150) were positive, including 125 (42% of 297) trace-positives. We recruited 109 of these 125 (87%) PWTS and additionally recruited 19 participants who were found to have trace-positive sputum during a national screening campaign^11^ to enroll a total of 128 PWTS (**Figure 2**). We also enrolled 139 matched Ultra-negative controls and 110 (76% of 144 eligible) positive controls.

**Figure 2.**
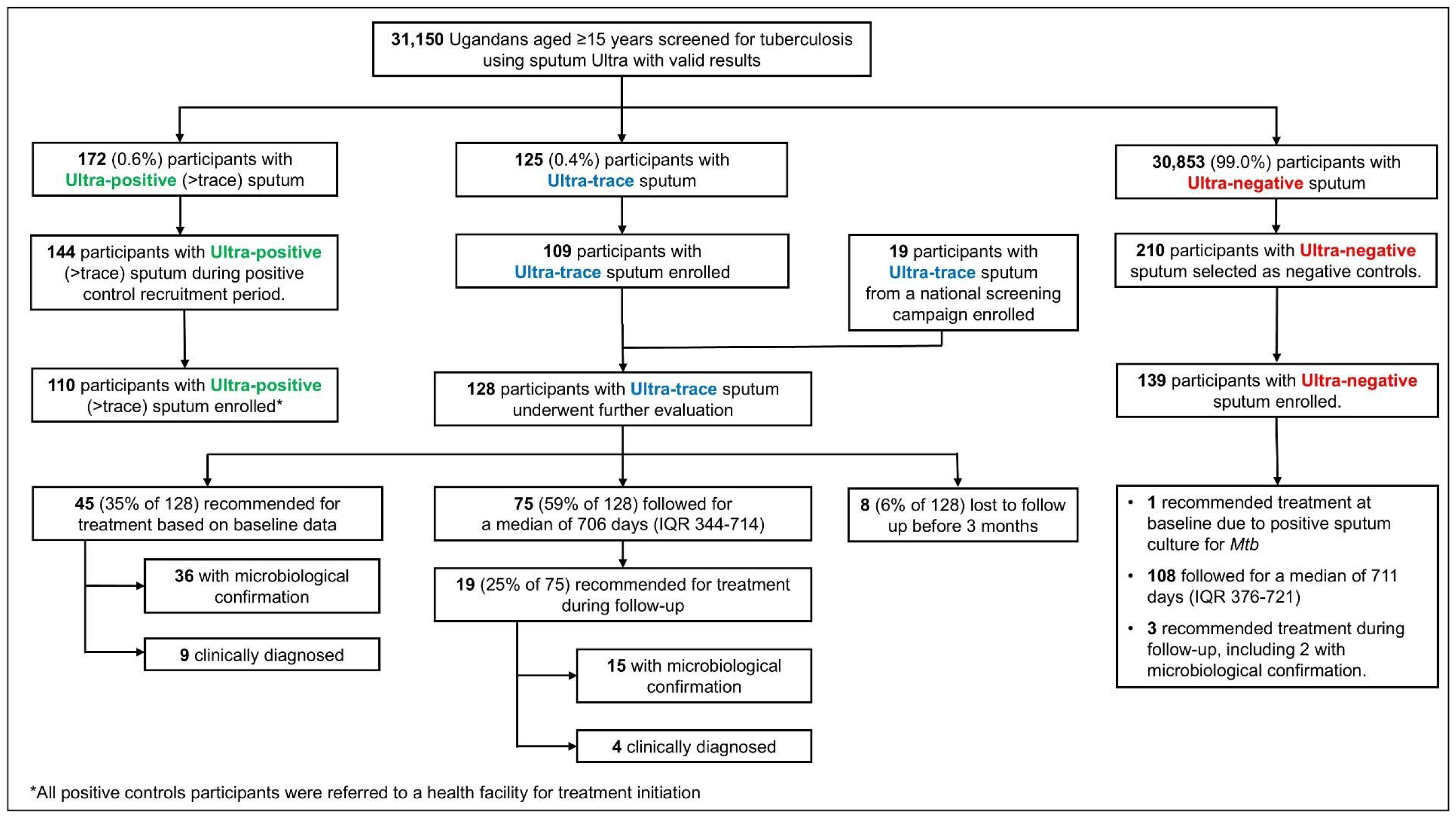
Recruitment and diagnostic outcomes of study participants with Ultra trace-positive sputum. The flow diagram illustrates the recruitment process and diagnostic outcomes of study participants. Most participants with trace-positive sputum were recruited from community-based tuberculosis screening conducted by the study team using Ultra as the initial test, and a minority of participants were recruited after undergoing similar symptom-neutral screening with Ultra through a national screening campaign. Of 498 participants (144, 144, and 210 with Ultra-positive, trace, and negative sputum results, respectively) evaluated for enrollment, 38 could not be contacted, 13 were ineligible, and 70 declined to participate (**appendix p 27**). Among participants with trace-positive sputum, there were 2 participants at baseline and 4 during follow-up with presumed false-positive microbiological results, for whom treatment was not recommended; these were not classified as “recommended for treatment” in this figure.

Among 128 PWTS, the median age was 33 (interquartile range [IQR]: 24-39), 62% (79/128) were male, 15% (19/128) had HIV infection, 46% (42/91 who were assessed) reported any cough at screening. (**Table 1**). Of 128 PWTS, 45 (35%) were recommended tuberculosis treatment based on additional testing performed at baseline, including 36 participants with microbiological confirmation and 27 with positive culture **(appendix pp 14-15)**. Of 45 PWTS recommended treatment at baseline, 34 had abnormal CXR, 41 had abnormal CT, and one did not complete imaging studies. Eight PWTS not recommended treatment were lost to follow-up before three months.

**Table 1.** Characteristics individuals screened for tuberculosis in community settings in Kampala, Uganda, according to initial sputum Ultra results and treatment recommendation.

|  | Positive Ultra (n=110) | Trace Ultra (n=121)<sup>#</sup> |  |  | Negative Ultra (n=139) |
| --- | --- | --- | --- | --- | --- |
|  |  | Treatment recommended at baseline (n=45) | Treatment recommended during follow-up (n=19) | Treatment not recommended<sup>#</sup> (n=56) |  |
| Age (years), median (IQR)<sup>†</sup> | 35 (28, 42) | 35 (24, 42) | 32 (27, 39) | 33.5 (25, 38) | 32 (25, 39) |
| Female, n (%)<sup>†</sup> | 26 (24%) | 13 (29%) | 7 (37%) | 25 (45%) | 60 (43%) |
| HIV positive, n (%)<sup>†</sup> | 23 (21%) | 9 (20%) | 4 (21%) | 6 (11%) | 11 (8%) |
| HIV positive, not on antiretroviral therapy at enrollment, n (%) | 4 (4%) | 1 (2%) | 2 (11%) | 0 (0%) | 1 (1%) |
| Household tuberculosis exposure within one year, n (%) | 15 (14%) | 7 (16%) | 4 (21%) | 7 (13%) | 7 (5%) |
| Screened during contact investigation, n (%) | 5 (5%) | 2 (4%) | 0 (0%) | 3 (5%) | 0 (0%) |
| Prior tuberculosis, n (%) | 20 (18%) | 4 (9%) | 7 (37%) | 12 (21%) | 6 (4%) |
| Prior tuberculosis within 2 years, n (%) | 13 (12%) | 3 (7%) | 3 (16%) | 5 (9%) | 3 (2%) |
| Current smoking, n (%) | 36 (33%) | 14 (31%) | 7 (37%) | 8 (14%) | 16 (12%) |
| Underweight (body mass index <18.5 kg/m<sup>2</sup>), n/N (%) | 27/102 (25%) | 17/45 (38%) | 2/18 (11%) | 4/46 (9%) | 11/125 (9%) |
| Cough at screening<sup>§</sup>, n/N (%) | 67/94 (71%) | 21/35 (60%) | 3/9 (33%) | 18/41 (44%) | 35/120 (29%) |
| Cough at enrollment, n (%) | 98 (89%) | 37 (82%) | 13 (68%) | 37 (66%) | 30 (22%) |
| Weeks of cough<sup>‡</sup>, median (IQR) | 5.5 (3, 16) | 4 (2, 8) | 2 (1, 6) | 3 (1, 12) | 2 (1, 4) |
| Cough severity on 0-100 visual analog scale, <sup>20,‡</sup> median (IQR) | 50 (30, 70) | 40 (20, 60) | 30 (10, 50) | 30 (20, 50) | 28 (10, 30) |
| Any tuberculosis symptom at screening<sup>*§</sup>, n (%) | 68/94 (72%) | 22/35 (63%) | 4/9 (44%) | 20/41 (49%) | 40/120 (33%) |
| Any tuberculosis symptom at enrollment<sup>**</sup>, n (%) | 104 (95%) | 40 (89%) | 17 (89%) | 47 (84%) | 58 (42%) |
| Positive QuantiFERON, n/N (%) | 59/67 (88%) | 35/40 (88%) | 12/15 (80%) | 32/53 (60%) | 51/106 (48%) |
| CRP ≥ 5.0 mg/L, n/N (%) | 56/103 (54%) | 19/44 (43%) | 4/18 (22%) | 13/56 (23%) | 21/136 (15%) |

<sup>†</sup> Matching constrained the age and sex of negative controls to be similar to, and HIV prevalence to be no greater than, that of PWTS as a whole. All participants were of black African ethnicity.
<sup>‡</sup> if cough reported.
<sup>\*</sup> any cough, fever, night sweats, or current weight loss at the time of screening
<sup>\*\*</sup> any cough, fever, night sweats, or weight loss (5kg over 12 month) at the time of enrollment
<sup>#</sup> This excludes 8 participants who completed less than 3 months of follow-up.
<sup>§</sup> at the time of initial screening, prior to receiving Ultra result; asked of all individuals screening after June 2021 HIV = human immunodeficiency virus, IQR = interquartile range, CRP = C-reactive protein

The cohort analyzed for tuberculosis diagnosis during follow-up consisted of 75 PWTS with no loss to follow-up before 3 months. They were followed for a median of 706 days (IQR 344–714), during which 19 (25% of 75) were recommended for tuberculosis treatment during follow-up, including 15 with microbiological confirmation (and 4 with microbiologically unconfirmed diagnoses [**appendix pp 16-17**], who were thus censored from the secondary analysis using a microbiologically-defined outcome). Therefore, when we considered diagnosis cumulatively from enrollment among all 120 PWTS with sufficient follow-up data for evaluation, 64 (53%, 95% CI 44-62%) were recommended for tuberculosis treatment either at baseline or during follow-up. Three deaths occurred among study participants, including one PWTS (**appendix p 13**).

Among PWTS not diagnosed with tuberculosis through baseline evaluation, the cumulative cause-specific hazard of tuberculosis diagnosis during follow-up was 0.24 (95% CI 0.15-0.40) at one year and 0.33 (95% CI 0.21-0.54) at two years when estimated based on treatment recommendation. Corresponding cumulative hazards based on microbiological positivity were 0.26 (95% CI 0.16-0.42) at one year and 0.36 (95% CI 0.22-0.58) at two years. The two-year cumulative hazard among Ultra-negative controls was 0.03 (0.01-0.10) based on treatment recommendation, or 0.02 (0.01-0.10) based on microbiological positivity (**Figure 3 a-b**). Results restricted to matched pairs were similar (**appendix p 18**).

**Figure 3.**
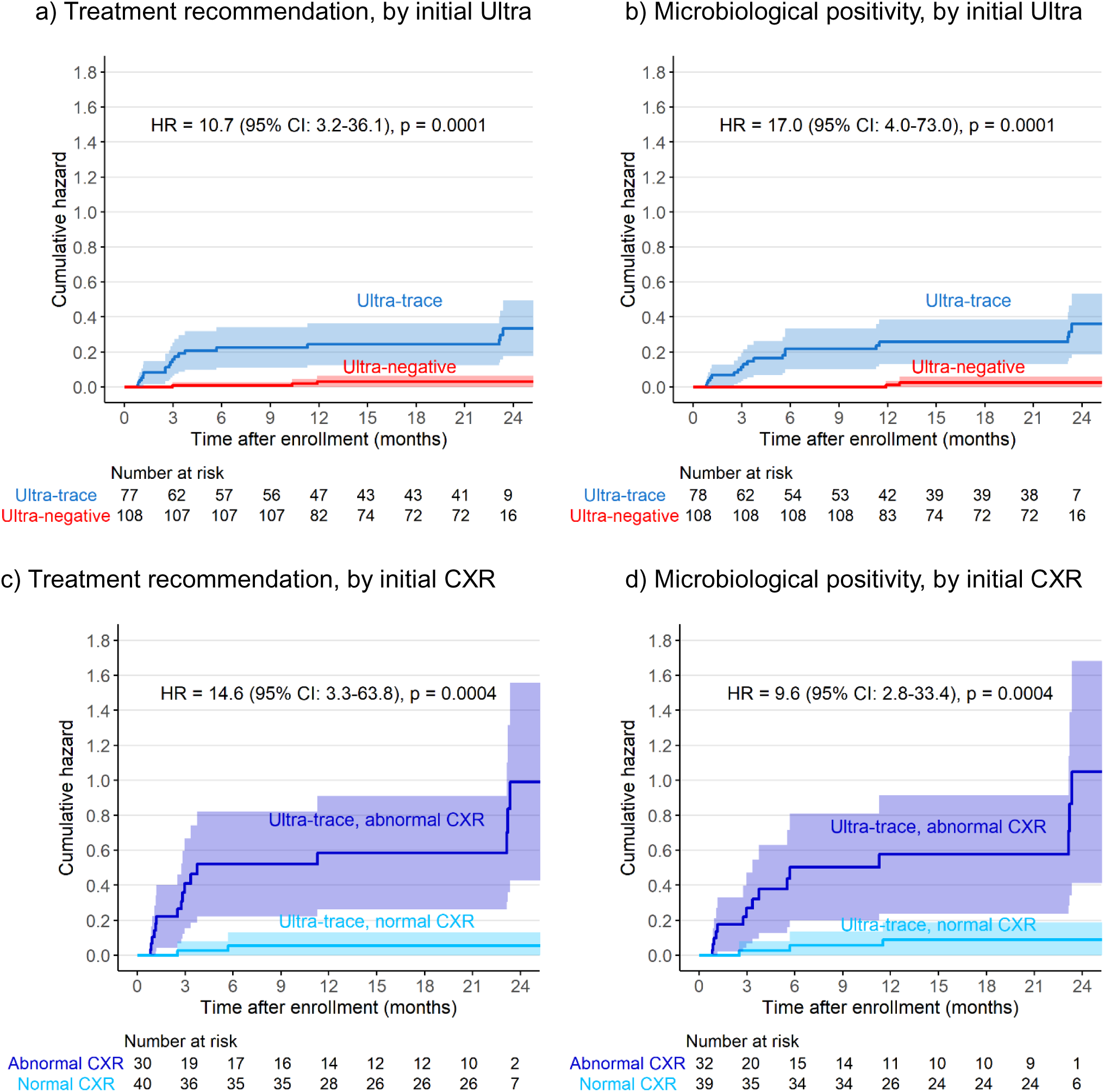
Cumulative cause-specific hazards of receiving a tuberculosis treatment recommendation (left) or developing a positive microbiological result for tuberculosis (right). The upper panels (a, b) present the cumulative hazard of tuberculosis diagnosis during follow-up among participants not diagnosed at baseline, estimated using the negative log transformation of Kaplan-Meier survival curves and stratified by participants’ initial Ultra results during community-wide tuberculosis screening. The lower panels (c, d) show results only for participants with initial trace-positive Ultra results, stratified by chest X-ray results at enrollment as interpreted by human readers. Shaded areas represent 95% confidence bands.

Among PWTS not diagnosed at baseline, tuberculosis diagnosis during follow-up was strongly associated with abnormal baseline CXR (HR 14.6 [95% CI 3.3-63.8, p=0.0004] as interpreted by human readers, **Figure 3 c-d**) but not with baseline cough (HR 1.2 95% CI 0.4-3.1, p=0.73) (**appendix p 19**). Sex, HIV status, and history of prior tuberculosis had elevated hazard ratios (>1) for tuberculosis diagnosis during follow-up but did not reach statistical significance in the primary analysis (**appendix p 22**).

The median CAD scores (by qXR v4) were highest among Ultra-positive controls (median 0.97 [IQR 0.86–0.99]) and lowest among negative controls (median 0.17 [IQR 0.06–0.36]). Among PWTS, those dignosed at baseline and during follow-up had similar median CAD scores (median 0.84 [IQR 0.52– 0.97] versus 0.87 [IQR 0.70–0.89]); PWTS followed for ≥3 months without tuberculosis diagnosis had lower CAD scores (median 0.30 [IQR 0.09–0.76]) than PWTS with tuberculosis diagnosis at baseline (p=0.0001) or during follow-up (p=0.055). Among PWTS not diagnosed with tuberculosis, elevated CAD scores occurred mainly in those with history of prior tuberculosis treatment (**Figure 4, Appendix p 23**).

**Figure 4.**
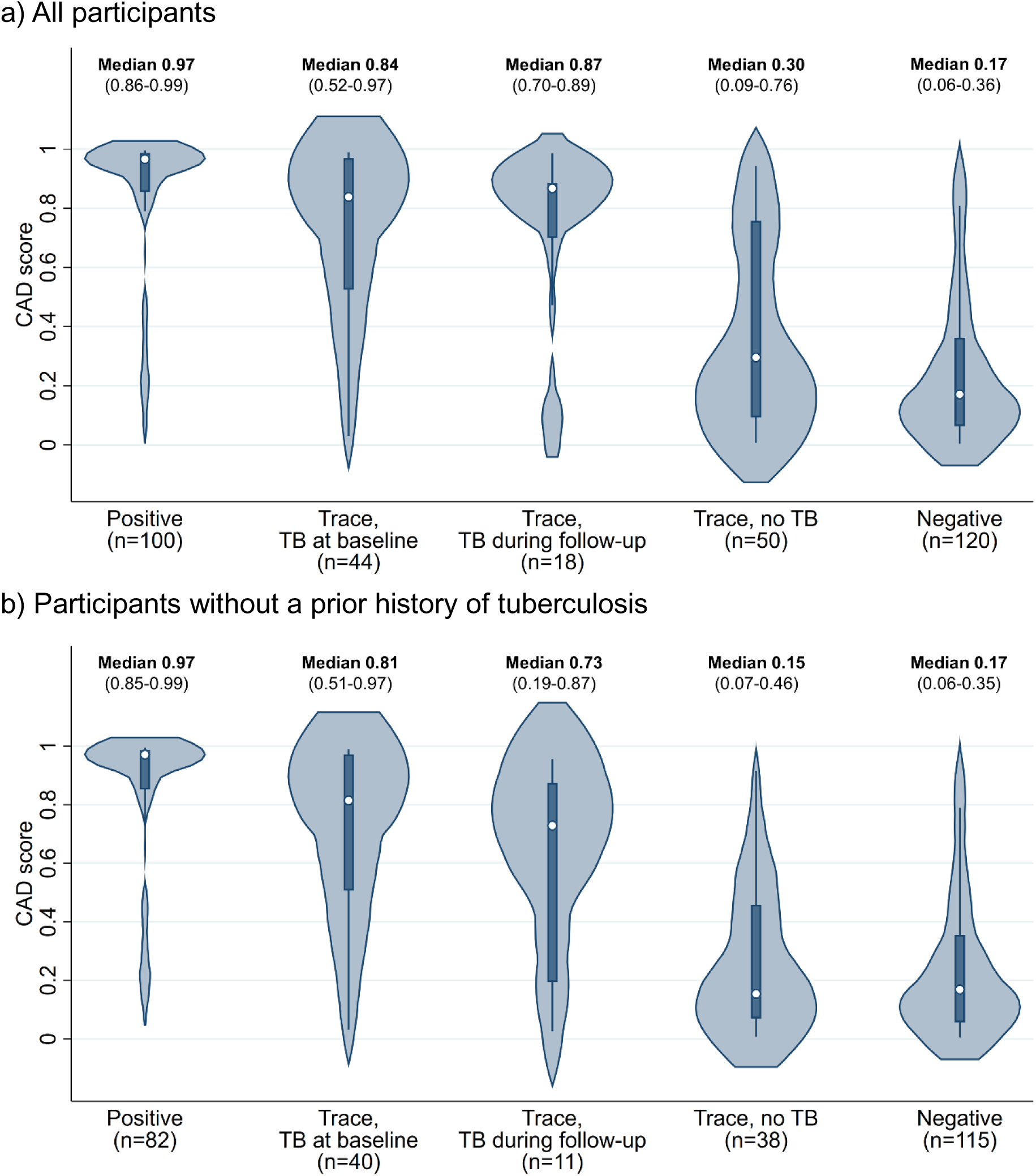
Distribution of CAD-interpreted baseline chest X-ray scores by initial sputum Ultra results and tuberculosis treatment recommendation. Results are shown for (a) all study participants and (b) participants without a prior history of tuberculosis, categorized into five groups: positive controls, PWTS diagnosed at baseline, PWTS diagnosed during follow-up, PWTS not diagnosed with TB, and negative controls. Curves represent the distribution of CAD scores based on kernel density estimates. Dots indicate median values, boxes show interquartile ranges, and whiskers extend to the upper and lower adjacent values for CAD scores in each group. The medians and interquartile ranges of CAD scores for each group are additionally described above each plot.

Baseline CXR interpretation by qXR v4 achieved an AUC of 0.77 (95% CI 0.69-0.86) for predicting tuberculosis at baseline or during follow-up among PWTS. AUC did not change when tuberculosis was defined microbiologically (AUC 0.77, 95% CI 0.68-0.86) (**Appendix p 24**). A baseline CAD score of 0.5 had a sensitivity of 77% (95% CI 66-86) for tuberculosis within 2 years (including baseline diagnoses) using a treatment-recommendation-based definition and 75% (62-84) using a microbiological definition, with 64% (50-76) specificity using either definition (**Appendix p 25)**. The estimates of the AUC were higher among PWTS without a prior history of tuberculosis treatment (AUC 0.84 [95% CI 0.76-0.92] versus 0.70 [0.47-0.92] for treatment recommendation [p=0.24]; AUC 0.80 [95% CI 0.70-0.89] versus 0.75 [0.53-0.97] for microbiological positivity [p=0.69]).

Of 62 PWTS recommended treatment at baseline or during follow-up who completed a CXR at baseline, only four (6%) had very low qXR scores (<0.1); all their baseline X-rays were also interpreted as normal by radiologists. One of them had a normal baseline chest CT but had untreated HIV and was started on tuberculosis treatment by her HIV clinician. The remaining three participants, all of whom had culture-confirmed tuberculosis, had subcentimeter nodules identifiable on their baseline chest CTs. Nearly all PWTS diagnosed with tuberculosis at baseline or during follow-up (60/63, 95%) had abnormal chest CTs at baseline (**Table 2**), compared to 36% (19/53) of PWTS who were not diagnosed.

**Table 2.** Baseline chest computed tomography characteristics of individuals screened for tuberculosis, according to initial sputum Ultra results and treatment recommendation.

|  | Positive Ultra (n=101) | Trace Ultra (n=117) |  |  | Negative Ultra (n=108) |
| --- | --- | --- | --- | --- | --- |
|  |  | Diagnosed at baseline (n=44) | Diagnosed during follow-up (n=19) | Not diagnosed (n=53) |  |
| **Any abnormality** | 96 (95%) | 41 (93%) | 19 (100%) | 19 (36%) | 24 (22%) |
| Consolidation | 55 (54%) | 20 (45%) | 4 (21%) | 4 (8%) | 3 (3%) |
| Ground glass opacity | 32 (32%) | 11 (25%) | 3 (16%) | 3 (6%) | 4 (4%) |
| Nodule ($\leq 3$cm) | 89 (88%) | 34 (77%) | 19 (100%) | 14 (26%) | 9 (8%) |
| Mass ($> 3$cm) | 3 (3%) | 1 (2%) | 0 (0%) | 0 (0%) | 0 (0%) |
| Septal thickening | 77 (76%) | 24 (55%) | 12 (63%) | 12 (23%) | 5 (5%) |
| Bronchial or bronchovascular abnormality | 79 (78%) | 25 (57%) | 10 (53%) | 12 (23%) | 7 (6%) |
| Cavitation | 53 (52%) | 10 (23%) | 2 (11%) | 2 (4%) | 1 (1%) |
| Parenchymal fibrosis | 61 (60%) | 23 (52%) | 11 (58%) | 15 (28%) | 7 (6%) |
| Volume loss | 34 (34%) | 11 (25%) | 5 (26%) | 7 (13%) | 1 (1%) |
| Emphysematous changes | 20 (20%) | 9 (20%) | 3 (16%) | 9 (17%) | 5 (5%) |
| Pleural thickening or fibrosis | 20 (20%) | 9 (20%) | 3 (16%) | 9 (15%) | 5 (5%) |
| Mediastinal or hilar lymphadenopathy | 14 (14%) | 6 (14%) | 3 (16%) | 0 (0%) | 0 (0%) |
| Cardiac abnormality | 0 (0%) | 0 (0%) | 0 (0%) | 1 (2%) | 2 (2%) |
| **Suggestive of current tuberculosis\*<sup>†</sup>** | 83 (82%) | 34 (77%) | 12 (63%) | 5 (9%) | 4 (4%) |
| **Suggestive of prior tuberculosis\*** | 59 (58%) | 26 (59%) | 13 (68%) | 13 (25%) | 7 (6%) |

\*Rated by a majority of radiologists as at least somewhat suggestive of current active or prior tuberculosis.
<sup>†</sup> Of baseline CTs interpreted as suggestive of current tuberculosis, 52/ 83 among positive controls, 23/34 among PWTS diagnosed at baseline, 9/12 among PWTS diagnosed during follow-up, and 5/5 among negative controls were also rated as suggestive of prior tuberculosis.

## Discussion

In this population-representative cohort of PWTS in a high-burden setting, the two-year risk of developing tuberculosis was high, even after a baseline evaluation that was interpreted as excluding tuberculosis disease. Approximately one-third were diagnosed through additional testing performed at enrollment, and of the remainder, the two-year cumulative hazard of tuberculosis diagnosis was 0.33. Among PWTS not diagnosed with tuberculosis at baseline, those with abnormal initial chest imaging had a particularly high risk of being diagnosed with tuberculosis during follow-up, whereas baseline symptoms showed no prognostic value.

Our report highlights that individuals who receive trace results from tuberculosis screening are at substantial risk of tuberculosis. While prior studies ^3–5, 7^ showed that 14% to 43% of screening participants with trace-positive sputum were culture-positive, our results demonstrate that culture-negative PWTS are still at significant risk of progressing to tuberculosis diagnosis over time. In this study, 53% of PWTS were diagnosed with tuberculosis within two years, and incompleteness of follow-up makes the true positive predictive value (PPV) even higher. Importantly, the PPV may be even higher in populations with a greater underlying prevalence of tuberculosis. For example, while this study used sputum Ultra as an initial test, most screening programs utilize Ultra as a confirmatory test among individuals with abnormal chest X-ray and/or symptoms.^3, 4^ Since the prevalence of tuberculosis is higher among those who screen positive, the PPV of the Ultra test will be also higher when used to confirm positive screening results.

When X-ray is used for screening, our results suggest that the screening step may also increase the probability that those with trace results have or will soon develop tuberculosis, even if further microbiological testing at the time of screening would be negative. Thus, our results support treatment for tuberculosis disease in most PWTS during tuberculosis screening, particularly when trace results are preceded by abnormal screening chest X-ray. The appropriate treatment strategy for this form of paucibacillary tuberculosis is uncertain, as lower-burden tuberculosis may be cured with shortened regimens.^15^

Our result that many PWTS may be culture negative yet at high risk to progress to more advanced disease — while some others could remain without disease over two years — can be explained by the current understanding of the tuberculosis disease spectrum and natural history. Tuberculosis has a dynamic disease course, and disease burden can fluctuate over time.^8, 10, 16^ Some culture-positive individuals may experience spontaneous improvement and become culture-negative.^9^ This immune-driven reduction in disease burden could leave nonviable *Mtb* that are detectable on molecular tests even after the burden of viable bacteria becomes too small for culture to detect ^17^. However, culture-negative individuals who once had enough mycobacterial burden to produce a positive Ultra result, and who have not received sterilizing treatment, could be at high risk of progressing to culture-positive disease The substantial proportion (nearly half) of PWTS with negative baseline cultures who were nevertheless diagnosed with tuberculosis during this study also suggests that the true specificity of Ultra in community screening contexts (which has already been shown^18^ to be higher than estimates in clinic-based populations) is even higher than previously estimated.

Our results also demonstrate the utility of chest imaging in interpreting Ultra trace results. More than half of PWTS without tuberculosis at baseline had CXRs interpreted as normal by radiologists, and this subset had a substantially lower risk of developing tuberculosis (cumulative hazard 0.05 at 2 years) — potentially too low to justify empiric treatment for tuberculosis disease, although still above the risk in contacts for whom preventive therapy is recommended.^19^ It should be noted that radiologists interpreting X-ray images in this study had access to paired CT images, and also that imaging results informed clinical decision-making. Thus, the estimated hazard ratio likely overestimated the ability of X-rays to discriminate early tuberculosis. However, X-ray scores by CAD software still demonstrated utility in identifying individuals diagnosed with tuberculosis among PWTS, particularly among those without prior tuberculosis.

This study has several limitations. Our screening using Ultra may have preferentially selected participants more capable of providing sputum, and PWTS at higher risk of tuberculosis may have been more likely to enroll into the study and complete follow-up. We did not induce sputum but accepted suboptimal quality samples when necessary, which may have underestimated microbiological positivity. Differential censoring of participants in the survival analysis may have biased our hazard estimates. In particular, those started on treatment without microbiological confirmation were censored from the analyses that used microbiological positivity as their outcome, but if untreated, they likely had a high risk of developing microbiological positivity later. Furthermore, if the highest-risk individuals developed tuberculosis early in follow-up, their absence from the cohort during later time periods may have biased our hazard ratio estimates. Under our primary definition of tuberculosis, clinical diagnoses may have led to overestimation of diagnosis at baseline, biased the cohort being followed toward lower-risk individuals, and/or overestimated two-year risk among that cohort. It is reassuring that our estimates were similar under our secondary, microbiological definition, although censoring individuals who started treatment and classifying false-positive microbiological results as positives may have led to under- or overestimation of hazards, respectively. The limited number of events in negative controls and PWTS with normal X-rays precluded adjustment for multiple covariates and resulted in wide confidence intervals. Lastly, our study was conducted in a single city and screening context, and results could differ in settings with different epidemiology of tuberculosis or different healthcare and care-seeking practices.

In summary, we conducted community-based tuberculosis screening using sputum Ultra in Kampala, and found that more than half of PWTS were confirmed to have tuberculosis within two years. Risk was elevated even among those not initially thought to have tuberculosis after extensive baseline diagnostic procedures. Tuberculosis diagnosis during follow-up was strongly associated with baseline chest imaging abnormalities but not with baseline symptoms. While further research is needed to determine the most appropriate treatment strategy for these individuals, our findings support tuberculosis treatment for most individuals with trace-positive screening results and abnormal chest X-rays. For individuals with negative microbiology and normal chest X-rays, individualized risk-benefit treatment decision making is needed, as their two-year risk of tuberculosis may still exceed that of contacts who are recommended for tuberculosis preventive treatment.

### Contributors

JS directly accessed and verified the underlying data, performed the data analysis and wrote the original draft of the manuscript. MN coordinated field data collection activities in Uganda. AN contributed to data management and data curation. PB and CV also contributed to data curation. JM, JA, RK, FK and MM enrolled participants and collected data in Uganda. CEK contributed to the laboratory processing of study samples. DWD contributed to the study’s conception and critically revised the manuscript. AK contributed to the study’s conception and supervised the data collection process in Uganda. EAK conceptualized the study, acquired funding and resources, supervised the data collection and development of the analytic plan, directly accessed and verified the underlying data, and critically revised the manuscript. All authors had full access to all the data in the study and accept responsibility for the decision to submit the manuscript for publication.

## Data sharing

The deidentified dataset used for this study and a data dictionary will be available upon a reasonable request. Data sharing will be limited to non-commercial research use only. Requests should include a proposal outlining the intended use and methodology and will be subject to review and approval. Proposals can be directed to.

## Acknowledgements

This work was supported by the National Institutes of Health (grant numbers R01HL153611 [to E. A. K.], R01HL138728 [to D. W. D.], T32AI007291 and K23AI185268 [to J.S.]) and Gates foundation (grant number INV-042921 to E.A.K.) The content is solely the responsibility of the authors and does not necessarily represent the official views of the funders.

