## supplementary appendix for "The long-term risk of tuberculosis among individuals with Xpert Ultra “trace” screening results: a longitudinal follow-up study"

### Supplementary Appendix Table of Contents

**Appendix A: Community-based tuberculosis screening and participant recruitment**

Screening was initially conducted in partnership with the STOMP-TB (Strategies for Treating, Observing, Managing, and Preventing Tuberculosis) study[1] from February 2021 through August 2021. During this period, screening was primarily door-to-door and took place in three contiguous parishes with an anticipated high burden of tuberculosis. From September 2021 to April 2024, screening was continued by the TURN-TB (Trace Ultra Result iNsight in TB screening) study[2] alone. This screening by TURN-TB was conducted throughout Kampala, in areas with similarly high anticipated TB burden. The TURN-TB study primarily employed event-based screening and selected locations based on TB notification rates from TB registers, as well as data on poverty and crowding.

In addition, participants with trace-positive sputum were also recruited from individuals who participated in similar community-based, symptom-agnostic Ultra testing offered by a national screening program (CAST-TB) in high-risk areas of Kampala.

**Appendix B: Selection of negative and positive control participants**

Negative controls were recruited from among the individuals screened with Ultra by the study, and were selected to be age- and sex-matched to participants with trace-positive sputum. We recruited negative controls of the same sex. For age, we applied a ±2-year caliper for individuals aged 54 or younger, and due to a more limited pool of older individuals, we used progressively broader age windows , increasing the caliper width by ±1 year for every 5-year increment in age beyond 55: ±3 years for ages 55–59, ±4 years for ages 60–64, ±5 years for ages 65–69, and so on. We also controlled the prevalence of HIV in negative control groups. Given that HIV status was not determined until after enrollment, we recruited an additional age- and sex-matched participant with Ultra-negative sputum each time that a negative control matched to an HIV-negative PWTS was found to be HIV positive, thus ensuring that each HIV-negative trace participant had an HIV-negative matched negative control.

The primary analysis is unmatched, as it excludes participants diagnosed with tuberculosis at baseline, resulting in the exclusion of one member of some matched pairs. We then additionally performed a sensitivity analysis, limiting the analysis to pairs in which both participants were not diagnosed at baseline and followed (results presented in Figure S1). For this analysis, we excluded any pair in which either the PWTS or the negative control was diagnosed at baseline or lacked follow-up data. For PWTS participants matched to more than one negative control (due to recruitment of an additional HIV-negative control after the first HIV-positive participant was enrolled), we included the first matched negative control in the survival analysis. If the first matched negative control lacked follow-up data, the second matched control was included instead

Positive controls were also recruited from among individuals screened with Ultra by the study. Consecutive participants with positive Ultra results (greater than trace) were enrolled until 110 participants were reached.

#### Appendix C. Sample size calculation for participants with trace-positive sputum

We powered our study to detect an elevated risk of incident TB among individuals with trace sputum that was similar to, or greater than, the risk among infected household contacts of an index patient with TB, for whom preventive therapy is recommended.[1] Among the participants in a prior active case finding study in Kampala,[2] the prevalence of Xpert-positive tuberculosis was 2-3 times the national adult average as estimated in a prevalence survey[3] and was higher still when using an event-based door screening approach. We therefore initially anticipated the incidence of tuberculosis among negative controls would be 0.5 to 1% per year, or 2.5 to 5 times the national estimated incidence,[4] translating to a 1 to 2% cumulative risk of TB diagnosis over two years. Based on earlier screening activities,[5] we estimated that 75% of individuals with trace-positive sputum would not have strong evidence of TB at baseline and could be followed for TB during follow-up. We estimated that enrolling 130 participants with trace-positive sputum would result in 80% power to distinguish a two-year cumulative risk of TB of 7-9% (3.5 to 4.5% per year) among individuals with trace-positive sputum compared to negative controls.

We powered our study to detect an elevated risk of incident TB among individuals with trace sputum that was similar or greater than the risk among infected household contacts of an index patient with TB, for whom preventive therapy is recommended.[1] Among the participants in a prior active case finding study in Kampala,[2] the prevalence of Xpert-positive tuberculosis was 2-3 times the national adult average as estimated in a prevalence survey[3] and was higher still when using an event-based door screening approach. Assuming the same relative incidence as relative prevalence, we initially anticipated the incidence of tuberculosis among negative controls would be 0.5 to 1% per year, or 2.5 to 5 times the national estimated incidence,[4] translating to 1 to 2% cumulative over two years. Based on earlier screening activities,[5] we estimated that 75% of individuals with trace-positive sputum would not have strong evidence of TB at baseline and could be followed up for incident TB. We estimated that enrolling 130 participants with trace-positive sputum would result in 80% power to detect a two-year cumulative TB incidence of 7-9% (3.5 to 4.5% per year) among individuals with trace-positive sputum as elevated compared to negative controls.

**Table S1: Minimum two-year cumulative incidence of tuberculosis among participants with trace-positive sputum required to achieve 80% power to detect a difference relative to negative controls.**

| | | **PWTS sample size required to reach 80% power** | | | | |
|---|---|---|---|---|---|---|
| | | **110** | **120** | **130** | **140** | **150** |
| **Two-year cumulative incidence of TB among negative controls** | **0.5%** | 6.2% | 5.8% | 5.5% | 5.3% | 5.0% |
| | **1.0%** | 7.7% | 7.3% | **6.9**% | 6.6% | 6.4% |
| | **2.0%** | 10.1% | 9.6% | **9.2%** | 8.9% | 8.6% |
| | **3.0%** | 12.1% | 11.6% | 11.2% | 10.8% | 10.4% |
| | **4.0%** | 13.9% | 13.4% | 12.9% | 12.5% | 12.2% |

References:
1. World Health Organization. WHO operational handbook on tuberculosis. Module 1: prevention-tuberculosis preventive treatment: World Health Organization; 2024.

2. Kendall EA, Kitonsa PJ, Nalutaaya A, Robsky KO, Erisa KC, Mukiibi J, et al. Decline in prevalence of tuberculosis following an intensive case finding campaign and the COVID-19 pandemic in an urban Ugandan community. Thorax. 2024;79(4):325-31.
3. The Republic of Uganda Ministry of Health. The Uganda National Tuberculosis Prevalence Survey, 2014-2015 Survey Report. 2016.
4. World Health Organization. Global Tuberculosis Report 2024. Geneva, Switzerland: WHO; 2024.
5. Kendall EA, Kitonsa PJ, Nalutaaya A, Erisa KC, Mukiibi J, Nakasolya O, et al. The Spectrum of Tuberculosis Disease in an Urban Ugandan Community and Its Health Facilities. Clin Infect Dis. 2021;72(12):e1035-e43.

**Appendix D. Symptom survey questionnaire at the time of enrollment**

| ***Questions*** | ***Response options*** |
|---|---|
| Which of the following symptoms do you have currently? (Currently can mean today or within the past few days.)<br>Select all that apply. | Cough<br>Coughing up blood<br>Unexplained fever or chills<br>Unexplained fatigue<br>Drenching sweats at night<br>Shortness of breath<br>Pain in my chest<br>Loss of normal appetite (Anorexia)<br>None of the above<br>Unknown/refused |
| Within the past twelve months, have you experienced weight loss of more than 5 kg, or enough to make your clothes loose? | No<br>Yes<br>Unknown/refused |
| Have you noticed any other signs or symptoms of illness that you are concerned about? | |
| What other symptoms of illness have you noticed? | |
| ***Conditional questions for participants who reported any symptoms.*** | |
| You said that you currently have a cough. Now looking back in time, for how long have you had this cough? (Interviewer should record answer in weeks.) | |
| You said that you are currently coughing up blood. Now looking back in time, for how long have you been coughing blood? (Interviewer should record answer in weeks.) | |
| You said that you currently have a fever or chills. Now looking back in time, for how long have you had fever or chills? (Interviewer should record answer in weeks.) | |
| You said that you currently have sweats at night. Now looking back in time, for how long have you had sweats at night? (Interviewer should record answer in weeks.) | |
| You said that you currently have unexplained fatigue. Now looking back in time, for how long have you had unexplained fatigue? (Record answer in weeks, rounding to the nearest week) | |
| You said that you currently have chest pain and/or shortness of breath. Now looking back in time, for how long have you had chest pain and/or shortness of breath?<br>(Record answer in weeks, rounding to the nearest week. If both symptoms are present, record the number of weeks for the symptom that has been present for the longest duration.) | |
| You said that you currently have lack of appetite. Now looking back in time, for how long have you had lack of appetite?<br>(Record answer in weeks, rounding to the nearest week) | |
| How long ago did you first notice your weight loss? (record answer in weeks) | |
| How do you think your weight is changing now? | I am continuing to lose weight |

<table>
<tr><td></td><td>My weight has stabilized<br>I am regaining or have regained weight<br>Unknown/Refused to answer</td></tr>
<tr><td>Use the visual below to indicate the severity of your cough in the past week. Please choose the point on this line that indicates the severity of your cough in the past week.<br>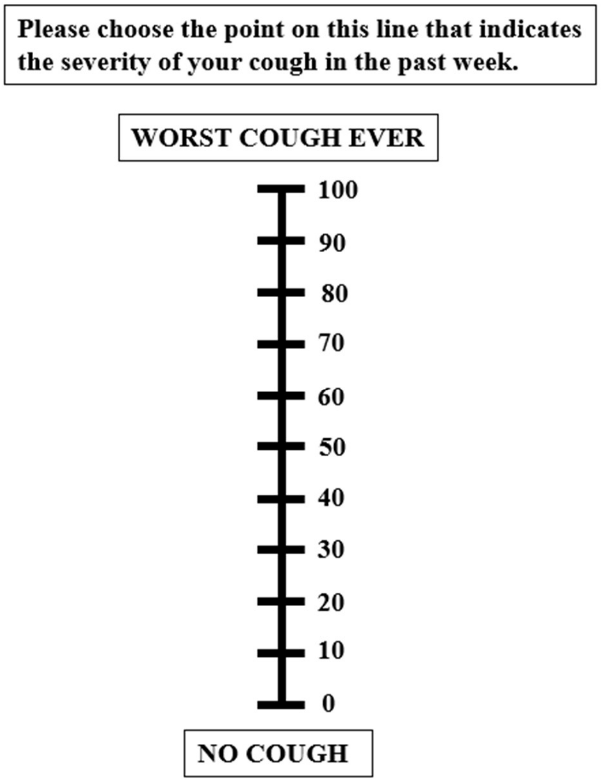<br></td><td></td></tr>
</table>

**Appendix E. Symptom survey questionnaire at the time of screening**

| ***Questions*** | ***Response options*** |
|---|---|
| TB sometimes has no symptoms, so we are testing everyone for TB regardless of whether or not they have symptoms. But we would like to know what symptoms you currently have.<br><br>Do you have cough? | No<br>Yes<br>Unable/unwilling to answer |
| For how many days have you had cough? | |
| Do you have sputum? | No<br>Yes<br>Unable/unwilling to answer |
| For how many days have you had sputum? | |
| Do you have blood stained sputum? | No<br>Yes<br>Unable/unwilling to answer |
| For how many days have you had blood-stained sputum? | |
| Do you have chest pain? | No<br>Yes<br>Unable/unwilling to answer |
| For how many days have you had chest pain? | |
| Do you have loss of body weight? | No<br>Yes<br>Unable/unwilling to answer |
| For how many days have you had loss of body weight?<br>*Approximate from when the participant thinks their weight loss began* | |
| Do you have fever? | No<br>Yes<br>Unable/unwilling to answer |
| For how many days have you had fever? | |
| Do you have excessive night sweats? | No<br>Yes<br>Unable/unwilling to answer |
| For how many days have you had excessive night sweats? | |

#### Appendix F. Radiologist interpretation and scoring system for chest imaging

Chest X-rays and CT scans from study participants were independently reviewed by two radiologists. If two interpretations were discrepant, a third independent radiologist conducted an additional review. All radiologists were blinded to clinical information and asked to assess the presence of specified abnormalities (e.g., nodules, cavities, fibrosis).

Radiologists were also asked to rate each study's consistency with current and (separately) prior tuberculosis using a four-point scale:

- **3**: Highly suggestive of (current or prior) tuberculosis
- **2**: Somewhat suggestive of (current or prior) tuberculosis
- **1**: Nonspecific finding that could be related to (current or prior) tuberculosis
- **0**: No radiographic evidence of (current or prior) tuberculosis

For each imaging study, we calculated the mean tuberculosis rating across all radiologists. Studies with a mean rating $\geq 1.5$ were classified as having imaging findings suggestive of (current or prior) tuberculosis. The 1.5 threshold was chosen to reflect the point at which at least half of the radiologists rated the imaging as at least somewhat suggestive of TB.

**Appendix G. Rationale for selecting definitions of tuberculosis**

Treatment recommendation was chosen as the primary definition of tuberculosis, as it allowed all evidence of tuberculosis status to be incorporated and was considered more likely to reflect the true disease state — including microbiologically unconfirmed disease — than microbiological positivity alone. Tuberculosis may be bacteriologically-negative (but apparent on imaging and by symptoms), and including expert clinicians' judgment in diagnosing these cases allowed us to avoid microbiological underdiagnosis, given that trace-positive tuberculosis is expected to be paucibacillary.[1] Conversely, a strictly microbiological definition classified six PWTS as having tuberculosis, whose results were not thought to represent true active disease when all available evidence was considered (including treatment history, clinical presentation, and discordance between different microbiological results). Clinicians therefore did not recommend treatment for these individuals, although they were classified as TB under the microbiological definition.

In the primary analysis, where treatment recommendation was used as the definition of tuberculosis, participants who were not recommended for tuberculosis treatment based on evaluations performed at baseline (i.e., upon enrollment) were included in the survival analysis. In the secondary analysis, where tuberculosis was defined exclusively by microbiological positivity, participants without positive microbiological testing results at baseline were included in the survival analysis.

**Appendix H. Time origins, endpoints, and competing events in survival analysis**

The origin and start of the survival analysis were both defined as the time when participants enrolled into the study, which typically occurred within one to two weeks (max 30 days) of screening. The end time was defined as the earliest occurrence of either tuberculosis diagnosis or death from causes other than tuberculosis; for participants who experienced neither event, the end time was the date of the last follow-up visit at which a sputum sample was obtained for microbiological testing.

In the primary analysis, which defined tuberculosis based on treatment recommendation, death from causes other than tuberculosis ("non-TB death") was considered a competing event for survival analysis. Among 77 PWTS who were not recommended for treatment at baseline and were followed for at least one month, no participants experienced a competing event (i.e., non-TB death). Among 108 negative controls who were not recommended treatment and followed for at least one month, one participant (1%) experienced a competing event (death during childbirth).

In the secondary analysis, which defined tuberculosis based on microbiological positivity, both non-TB death and initiation of tuberculosis treatment without microbiological confirmation were considered competing events for survival analysis. Among 78 PWTS without microbiological positivity at baseline and followed for at least one month, no participants experienced non-TB death, but 12 (15%) initiated treatment without microbiological confirmation before completing study follow-up. Among 108 negative controls without microbiological positivity at baseline and followed for at least one month, one participant (1%) experienced non-TB death, and one participant (1%) initiated tuberculosis treatment without microbiological confirmation prior to completing follow-up.

Participants who experienced a competing event were treated as censored in each analysis.

**Appendix I: Safety considerations and procedures for participants enrolled within two years of study completion**

Three deaths occurred among study participants: One PWTS living with HIV died of tuberculosis at 12 months after missing their six-month study visit and stopping antiretroviral treatment; one negative-control participant died during childbirth; and a positive-control participant died shortly after enrollment, before initiating treatment.

Study progress was monitored semi-annually by an Observational Study Monitoring Board (OSMB). In June 2024, based on preliminary findings, the OSMB recommended that participants with initial trace screening results, no history of prior tuberculosis treatment, and no tuberculosis diagnosis during the study be referred for tuberculosis preventive treatment when they completed study follow-up, and these recommendations were implemented accordingly.

Study follow up ended twelve months after enrolling the last participant. Therefore, not all participants were able to complete a full 24 months of follow-up before the end of the study. All participants had sufficient time to complete 12-month follow-up visits, but 23% (62 out of 267) of participants with initial trace or negative screening results were enrolled less than 24 months before the study ended. For individuals with trace screening results who were still in follow-up at the close of the study, a study visit was conducted in the final month of the study for those enrolled 16-23 months earlier, whereas follow-up time was assumed to conclude at the date of most recent visit for those enrolled ≤15 months earlier.

**Table S2: Participants with trace-positive sputum results at screening who had negative microbiological work up but were recommended TB treatment at baseline.**

| Age range, years old | Sex | HIV status | Prior TB | BMI, kg/m$^2$ | TB symptoms at enrollment | Baseline IGRA | Baseline CRP | qXR TB score from baseline CXR* | Baseline CT chest | Rationale for clinical diagnosis at baseline |
|---|---|---|---|---|---|---|---|---|---|---|
| 26-30 | M | Neg | No | 17.8 | Fever and night sweats for 12 weeks. Weight loss for 4 weeks. No cough. | Not done | <2.5mg/L | 0.81 | Multiple nodules in upper lobes, and a small cavity seen in the apical segment of the right upper lobe. Multifocal areas of patchy consolidation in the left upper lobe. | Constitutional symptoms, low BMI, CT findings highly consistent with active TB |
| 21-25 | F | Pos | No | 22.3 | Cough for 2 weeks | Negative | <2.5mg/L | 0.03 | Unremarkable | Started on treatment by a non-study clinician based on clinical risk (positive HIV and not on ART) |
| 21-25 | F | Neg | No | 25.9 | None | Positive | <2.5mg/L | 0.41 | A large, thick-walled cavitary nodule, along with a few additional nodules in the right upper lobe. | Baseline CT highly suggestive of active tuberculosis |
| 36-40 | M | Neg | No | 20.3 | None | Indeterminate | 6.05mg/L | 0.67 | Bilateral fibrotic and nodular infiltrates. | Repeat sputum Xpert trace-positive and CT concerning for active TB |
| 41-45 | M | Neg | Yes | 19.8 | Cough for 16 weeks, fever/chills for 8 weeks, weight loss for 28 weeks, shortness of breath and chest pain/shortness of breath for 28 weeks | Positive | 7.35mg/L | 0.79 | Extensive parenchymal fibrosis and bronchiectasis throughout both lungs, multiple nodules and cavities. | Started on treatment by a non-study clinician, based on prior tuberculosis with incomplete treatment. At enrollment the participant reported a history of tuberculosis treatment over a year ago, but he was later found to have not completed the prior course of treatment. |

| | | | | | | | | | | |
|---|---|---|---|---|---|---|---|---|---|---|
| 26-30 | M | Neg | No | 17.6 | Cough, weight loss, and fatigue for 2 weeks; shortness of breath and chest pain for 7 weeks. | Positive | <2.5mg/L | 0.98 | Two mass like opacities in the right lung apex, surrounded by multiple solid nodules. | Started on treatment by a non-study clinician, after the participant presented to a health facility with persistent cough and body weakness. |
| 36-40 | M | Pos | No | 20.7 | Cough, hemoptysis, fatigue, shortness of breath, and chest pain for 2 weeks, and weight loss for 1 week. | Negative | 11.49mg/L | 0.49 | A few random sub-centimeter solid nodules scattered in both lungs | Started on treatment by a non-study clinician, given a new HIV diagnosis. |
| 31-35 | M | Neg | No | 19.5 | Cough for 3 weeks, fever and chills for 1 week, and weight loss for 4 weeks | Positive | <2.5mg/L | 0.99 | Multiple centrilobular nodules in both upper lobes, with a small thick-walled cavity is seen in the right upper lobe. Patchy areas of consolidation around the nodules. | Recent TB contact, symptoms, and imaging consistent with active TB. |
| 21-25 | M | Neg | No | 19.0 | Cough for 4 weeks, weight loss for 15 weeks | Positive | 5.01mg/L | 0.82 | Multiple tiny centrilobular nodules. Patchy consolidation in the apical segment of the right upper lobe. | Recent TB household contact, symptoms, and CT suggestive of early active TB |

*qXR TB scores were not available to clinicians making treatment decisions, but they could review chest images.

**Table S3: Participants with trace-positive sputum at screening who subsequently had only negative microbiological results but were recommended TB treatment during follow-up.**

| Age range, years old | Sex | HIV status | Prior TB | BMI, kg/m² | TB symptoms at enrollment | Baseline IGRA | Baseline CRP ≥ 5.0mg/L | qXR TB score from baseline CXR* | Baseline CT chest | Rationale for clinical diagnosis during follow-up. |
|---|---|---|---|---|---|---|---|---|---|---|
| 41-45 | M | Neg | No | 21.3 | Cough for 2 weeks, fever/chills for 4 weeks, night sweats for 12 weeks, and chest pain/shortness of breath for 12 weeks | Indeterminate | No | 0.88 | Multiple solid nodules in the right upper lobe, including one containing a small cavity. The smaller nodules form branching tree-in-bud opacities. | Treatment was recommended at 3 months, based on a repeat CT which showed progressive peribronchial consolidation with associated centrilobular tree-in-bud nodules in the right upper lobe and increasing ipsilateral hilar lymphadenopathy. |
| 21-25 | F | Neg | No | 18.8 | Cough and weight loss for 26 weeks, night sweats for 2 weeks, and cough/shortness of breath for 16 weeks | Positive | No | 0.82 | A solid lobulated solitary nodule in the left upper lobe associated with ipsilateral hilar lymphadenopathy. | Treatment was recommended at 3 months, based on persistent symptoms, positive baseline IGRA, and a repeat CT scan at 3 months showing a left upper lobe nodule that had decreased in size but remained present. |
| 26-30 | F | Pos | Yes | 15.7 | Cough and chest pain/shortness of breath for 8 weeks, fever/chills for 4 weeks, night sweats for 8 weeks, and weight loss for 52 weeks. | Negative | Yes | 0.87 | Diffuse fibrosis and bronchiectasis in both upper lobes | Started on treatment by a non-study clinician, after discovering that the participant had been diagnosed with Xpert+ TB ~5 months before enrollment and completed only 1 month of treatment. |
| 51-55 | M | Neg | Yes | 19.5 | Cough for 6 weeks, | Positive | Yes | 0.92 | Right upper lobe fibrosis. A few random | Started on treatment by a non-study clinician one |

| | | | | | | | | | | |
|---|---|---|---|---|---|---|---|---|---|---|
| | | | | | fever/chills for 3 weeks, night sweats for 6 weeks, weight loss for 2 weeks, and joint pain. | | | | solid nodules in the left lower lobe. | month after study enrollment, due to persistent symptoms and a history of prior incomplete (~4 months) TB treatment four years ago. |

*qXR TB scores were not available to clinicians making treatment decisions, but they could review chest images.

**Figure S1. Cumulative cause-specific hazards of receiving a tuberculosis treatment recommendation (left) or developing a positive microbiological result for tuberculosis (right), among age- and sex-matched pairs of participants with Ultra trace versus negative screening results.** All PWTS and negative controls were matched on enrollment, and this analysis excluded any pair in which either the PWTS or the negative control was diagnosed at baseline or lacked follow-up data. The upper panels (a, b) present the cumulative hazard of tuberculosis diagnosis during follow-up among participants not diagnosed at baseline, estimated using the negative log transformation of Kaplan-Meier survival curves and stratified by participants' initial Ultra results during community-wide tuberculosis screening. The lower panels (c, d) show results only for participants with initial trace-positive Ultra results, stratified by chest X-ray results at enrollment as interpreted by human readers. Shaded areas represent 95% confidence intervals.

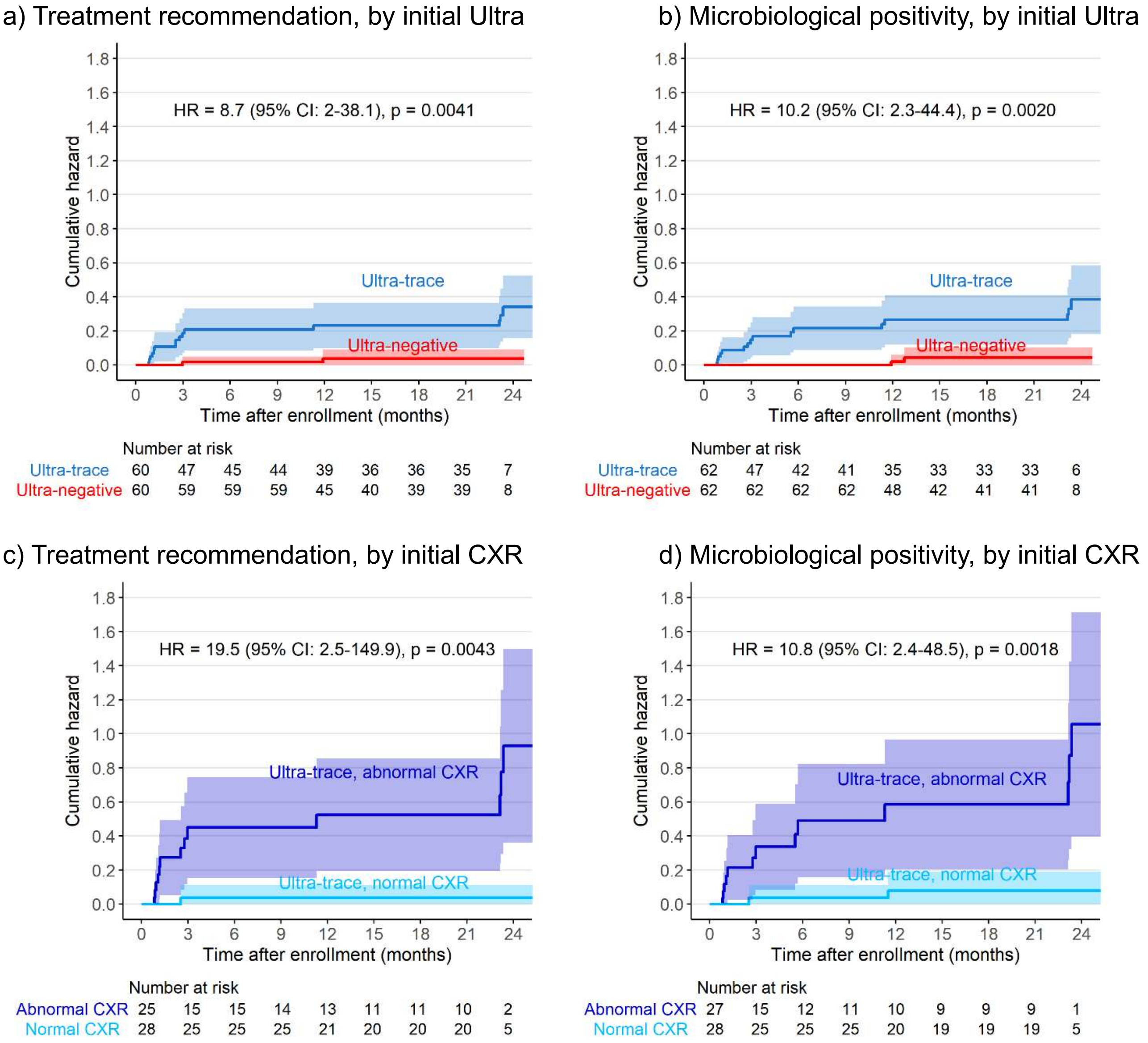

**Figure S2. Cumulative cause-specific hazards of receiving tuberculosis treatment recommendation (left) or developing a positive microbiological result (right) among individuals with trace screening results, stratified by whether cough was reported at the time of enrollment (top row) or at the time of screening (bottom row).** Abbreviations: HR (hazard ratio); CI (confidence interval); CXR (Chest X-ray)

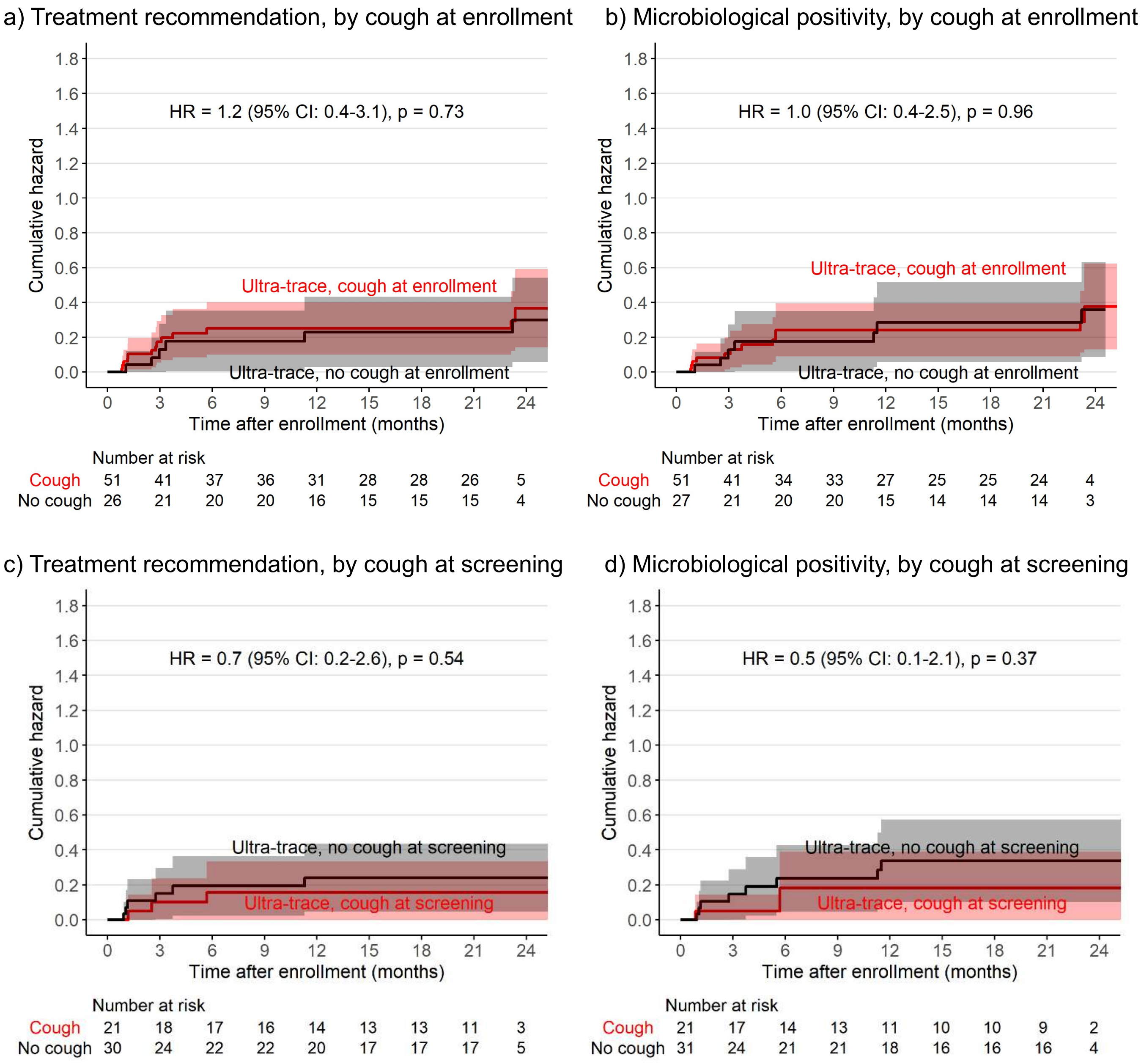

**Figure S3. Cumulative cause-specific hazards of receiving tuberculosis treatment recommendation (left) or developing a positive microbiological result (right) among individuals with trace screening results, stratified by baseline chest X-ray results interpreted by computer-aided detection software (qXR v4).** Abbreviations: HR (hazard ratio); CI (confidence interval); CXR (Chest X-ray)

a) Treatment recommendation, by CAD threshold of 0.2

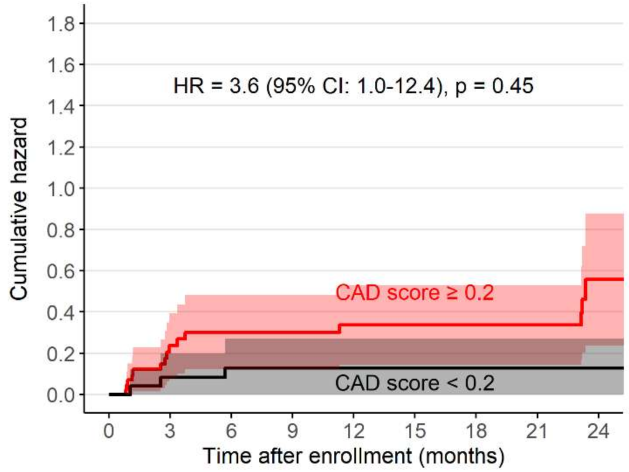

Number at risk

| | | | | | | | | | |
|---|---|---|---|---|---|---|---|---|---|
| Score≥0.2 | 45 | 32 | 30 | 29 | 23 | 20 | 20 | 18 | 4 |
| Score<0.2 | 25 | 23 | 22 | 22 | 19 | 18 | 18 | 18 | 5 |

b) Microbiological positivity, by CAD threshold of 0.2

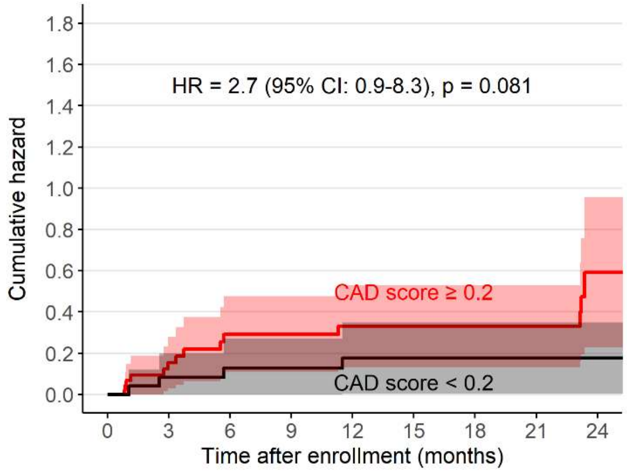

Number at risk

| | | | | | | | | | |
|---|---|---|---|---|---|---|---|---|---|
| Score≥0.2 | 46 | 32 | 27 | 26 | 19 | 17 | 17 | 16 | 2 |
| Score<0.2 | 25 | 23 | 22 | 22 | 18 | 17 | 17 | 17 | 5 |

a) Treatment recommendation, by CAD threshold of 0.5

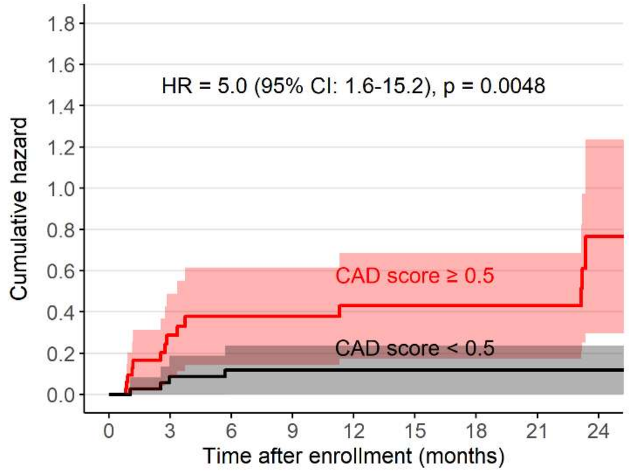

Number at risk

| | | | | | | | | | |
|---|---|---|---|---|---|---|---|---|---|
| Score≥0.5 | 33 | 23 | 21 | 20 | 17 | 15 | 15 | 13 | 3 |
| Score<0.5 | 37 | 32 | 31 | 31 | 25 | 23 | 23 | 23 | 6 |

b) Microbiological positivity, by CAD threshold of 0.5

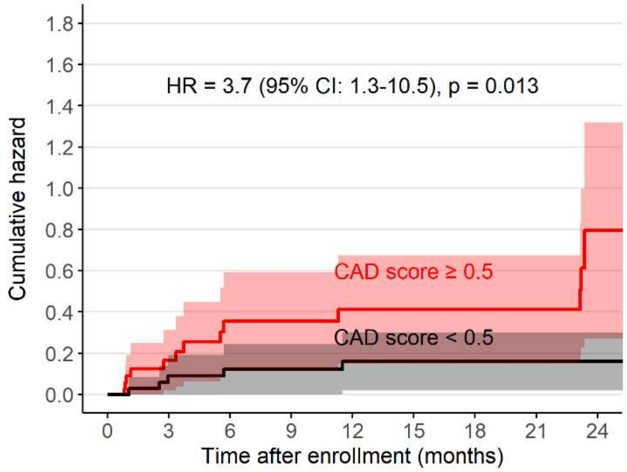

Number at risk

| | | | | | | | | | |
|---|---|---|---|---|---|---|---|---|---|
| Score≥0.5 | 35 | 24 | 19 | 18 | 14 | 13 | 13 | 12 | 2 |
| Score<0.5 | 36 | 31 | 30 | 30 | 23 | 21 | 21 | 21 | 5 |

**Figure S4. Cumulative cause-specific hazards of receiving a tuberculosis treatment recommendation (left) or developing a positive microbiological result for tuberculosis (right), among tuberculosis treatment-naïve participants with trace-positive screening results.** These figures show the cumulative hazard of tuberculosis diagnosis during follow-up among participants with an initial trace-positive Ultra result and no prior history of tuberculosis treatment, stratified by chest X-ray findings at enrollment as interpreted by human readers. Shaded areas represent 95% confidence intervals.

a) Treatment recommendation, by initial CXR

b) Microbiological positivity, by initial CXR

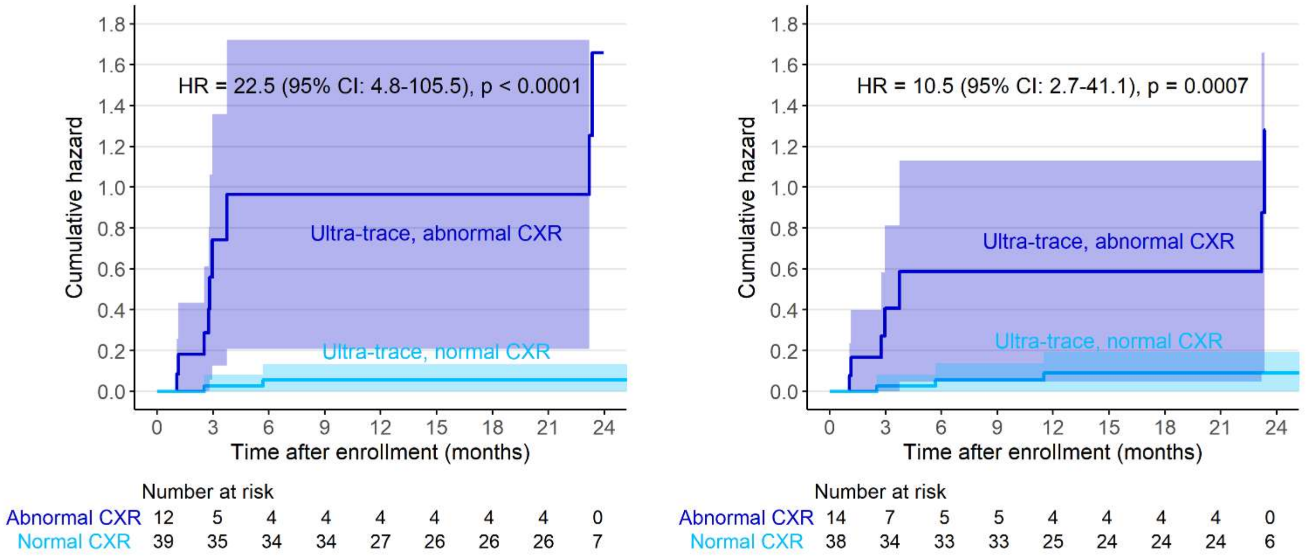

**Table S4a. Association between individual characteristics and tuberculosis diagnosis during follow-up, using a treatment-recommendation-based definition of tuberculosis.**

| | Events in subgroup, total n/N=20/78 (26%) | Hazard Ratio | 95% CI | p-value |
|---|---|---|---|---|
| **Male sex** | 12/44 (27%) | 1.4 | 0.6-3.6 | 0.4775 |
| **HIV infection** | 4/10 (40%) | 1.9 | 0.6-5.8 | 0.2512 |
| **Cough on enrollment** | 13/51 (25%) | 1.2 | 0.4-3.1 | 0.7338 |
| **Any tuberculosis symptom on enrollment** | 17/65 (26%) | 1.6 | 0.4-6.8 | 0.5529 |
| **History of prior TB** | 7/19 (37%) | 2.1 | 0.8-5.4 | 0.1150 |
| **CXR suggestive of active tuberculosis*** | 5/11 (45%) | 2.0 | 0.7-5.6 | 0.1867 |
| **CXR with CAD score ≥0.2** | 15/45 (33%) | 3.6 | 1.0-12.4 | 0.0445 |
| **CXR with CAD score ≥0.5** | 14/33 (42%) | 5.0 | 1.6-15.2 | 0.0048 |
| **CXR with any abnormality*** | 16/30 (53%) | 14.6 | 3.3-63.8 | 0.0004 |
| **CT suggestive of active tuberculosis*** | 12/17 (71%) | 7.7 | 3.0-19.5 | <0.0001 |

**Imaging results at baseline evaluation*

**Table S4b. Association between individual characteristics and tuberculosis diagnosis during follow-up, using a microbiological positivity-based definition of tuberculosis.**

| | Events in subgroup, total n/N=20/79 (25%) | Hazard Ratio | 95% CI | p-value |
|---|---|---|---|---|
| **Male sex** | 13/45 (29%) | 1.9 | 0.7-5.1 | 0.1879 |
| **HIV infection** | 4/10 (40%) | 2.0 | 0.6-6.0 | 0.2311 |
| **Cough on enrollment** | 12/51 (24%) | 1.0 | 0.4-2.5 | 0.9644 |
| **Any tuberculosis symptom on enrollment** | 17/65 (26%) | 1.8 | 0.4-7.9 | 0.4268 |
| **History of prior TB** | 8/19 (42%) | 3.0 | 1.2-7.5 | 0.0200 |
| **CXR suggestive of active tuberculosis*** | 8/13 (62%) | 4.8 | 1.9-12.3 | 0.0010 |
| **CXR with CAD score ≥0.2** | 14/46 (30%) | 2.7 | 0.9-8.3 | 0.081 |
| **CXR with CAD score ≥0.5** | 13/35 (37%) | 3.7 | 1.3-10.5 | 0.0126 |
| **CXR with any abnormality*** | 15/32 (47%) | 9.6 | 2.8-33.4 | 0.0004 |
| **CT suggestive of active tuberculosis*** | 13/20 (65%) | 9.5 | 3.6-25.2 | <0.0001 |

**Imaging results at baseline evaluation*

**Figure S5. Distribution of CAD-interpreted baseline chest X-ray scores by initial sputum Ultra results and tuberculosis treatment recommendation, presented as box-and-whisker plots.** Results are shown for (a) all study participants and (b) participants without a prior history of tuberculosis, categorized into five groups: positive controls, PWTS diagnosed at baseline, PWTS diagnosed during follow-up, PWTS not diagnosed with TB, and negative controls. Each box represents the interquartile range (IQR), with the horizontal line indicating the median CAD score. Whiskers extend to the most extreme values within 1.5 times the IQR from the lower and upper quartiles. Values beyond this range are shown as individual dots and represent outliers.

a) All participants

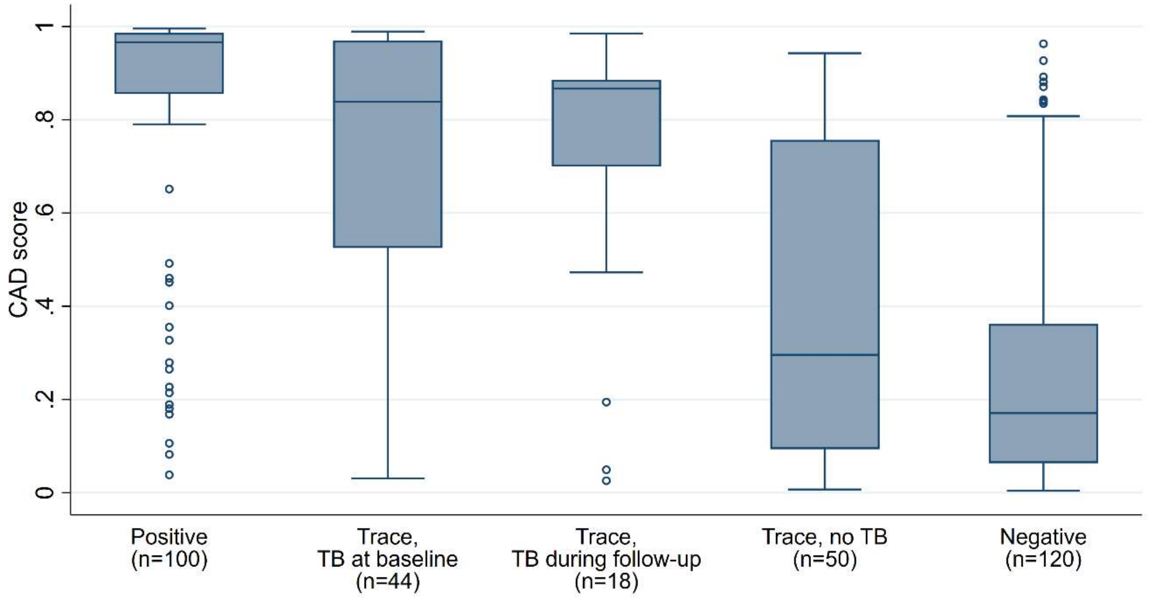

b) Participants without a prior history of tuberculosis

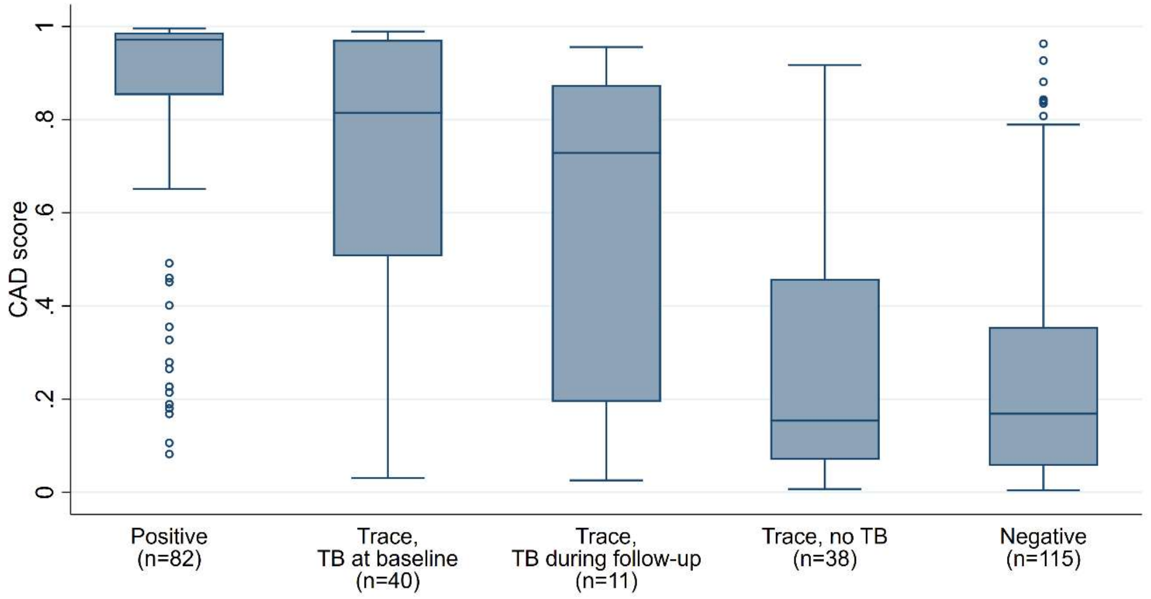

**Figure S6. Receiver operating characteristic (ROC) curves of computer-aided detection software (qXR v4) interpretations of baseline chest X-rays for predicting tuberculosis disease.** The reference standard for these plots is a treatment recommendation (left panels) or microbiological positivity (right panels), among all individuals with trace-positive screening results (a, b) or only those with no prior history of tuberculosis (c, d). This analysis included participants who completed chest X-rays at enrollment and were either diagnosed with tuberculosis (at baseline or during follow-up) or followed for at least 3 months without a tuberculosis diagnosis. Abbreviations: AUC (area under the curve); CI (confidence interval)

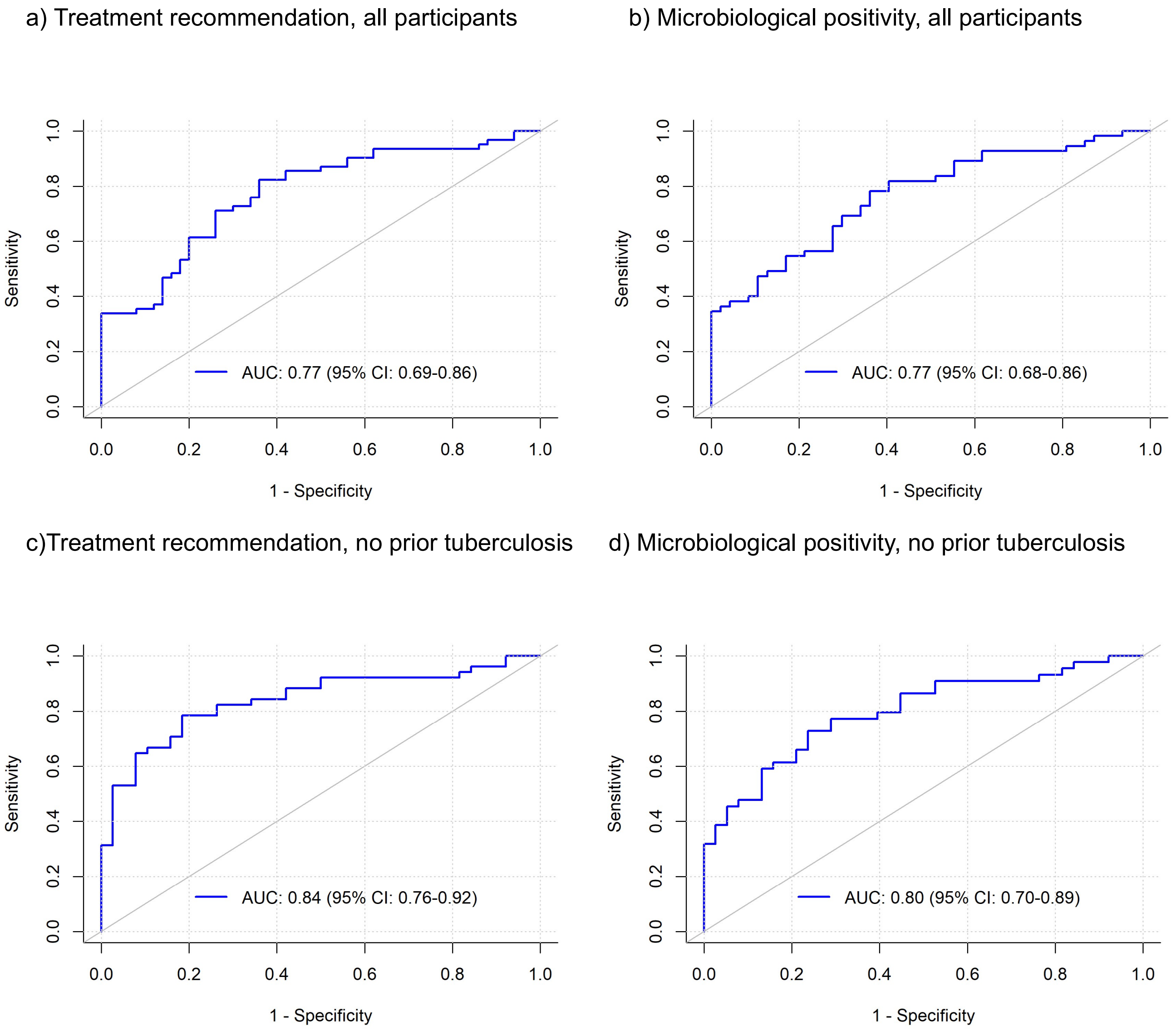

**Table S5a. Sensitivities and specificities of a computer-aided detection software (qXR v4) interpretations of baseline chest X-rays for predicting tuberculosis disease (either at baseline or during follow-up) among individuals with trace-positive screening results.**

| qXR threshold | Treatment recommendation | | Microbiological positivity | |
|---|---|---|---|---|
| | Sensitivity (95% CI) | Specificity (95% CI) | Sensitivity (95% CI) | Specificity (95% CI) |
| **0.1** | 94% (85-97) | 26.0% (16-40) | 93% (83-97) | 26% (15-40) |
| **0.2** | 87% (77-93) | 44% (31-58) | 85% (74-92) | 45% (31-59) |
| **0.3** | 87% (77-93) | 50% (37-63) | 84% (72-91) | 49% (35-63) |
| **0.4** | 85% (75-92) | 56% (42-69) | 82% (70-90) | 55% (41-69) |
| **0.5** | 77% (66-86) | 64% (50-76) | 75% (62-84) | 64% (50-76) |
| **0.6** | 71% (59-81) | 72% (58-83) | 65% (52-77) | 70% (56-81) |
| **0.7** | 66% (54-77) | 74% (60-84) | 62% (49-73) | 72% (58-83) |
| **0.8** | 58% (46-70) | 80% (67-89) | 55% (41.5-67) | 79% (65-88) |
| **0.9** | 35% (25-48) | 88% (76-94) | 38% (27-51) | 91% (80-97) |

**Table S5b. Sensitivities and specificities of a computer-aided detection software (qXR v4) interpretations of baseline chest X-rays for predicting tuberculosis disease (either at baseline or during follow-up) among individuals with trace-positive screening results and no history of prior tuberculosis.**

| qXR threshold | Treatment recommendation | | Microbiological positivity | |
|---|---|---|---|---|
| | Sensitivity | Specificity | Sensitivity | Specificity |
| **0.1** | 92% (82-97) | 34% (21-50) | 91% (79-96) | 32% (19-47) |
| **0.2** | 84% (72-92) | 58% (42-72) | 82% (68-90) | 55% (40-70) |
| **0.3** | 84% (72-92) | 66% (50-79) | 80% (65-89) | 61% (45-74) |
| **0.4** | 82% (70-90) | 71% (55-83) | 77% (63-87) | 66% (50-79) |
| **0.5** | 73% (59-83) | 82% (67-91) | 68% (53-80) | 76% (61-87) |
| **0.6** | 65% (51-76) | 89% (76-96) | 59% (44-72) | 84% (70-93) |
| **0.7** | 59% (45-71) | 92% (79-97) | 55% (40-68) | 87% (73-94) |
| **0.8** | 51% (38-64) | 97% (87-100) | 45% (32-60) | 92% (79-97) |
| **0.9** | 31% (20-45) | 97% (87-100) | 32% (20-47) | 97% (87-100) |

**Table S6: Number of participants evaluated for enrollment in each arm and their enrollment outcomes**

| Screening Ultra result | Positive | Trace | Negative |
|---|---|---|---|
| Initially selected for enrollment, n | 144 | 144<br>(125 from a study-conducted screening and 19 from a national screening) | 210 |
| Not able to be contacted | 8 | 7 | 23 |
| Ineligible, n | 9<br>• Already on treatment (n=3)<br>• Communication barrier (n=2)<br>• Residence outside the study follow-up area (n=1)<br>• Duplicate screening after enrollment (n=3). | 0 | 4<br>• Communication barrier (n=2)<br>• Residence outside the study follow-up area (n=1)<br>• Not appropriately age-matched to a participant with trace-positive sputum (n=1) |
| Declined to participate, n | 17 | 9 | 44 |
| Consenting and enrolled, n | 110 | 128 | 139 |
